# Evaluation of four commercial, fully automated SARS-CoV-2 antibody tests suggests a revision of the Siemens SARS-CoV-2 IgG assay

**DOI:** 10.1101/2020.11.27.20239590

**Authors:** Christian Irsara, Alexander E. Egger, Wolfgang Prokop, Manfred Nairz, Lorin Loacker, Sabina Sahanic, Thomas Sonnweber, Wolfgang Mayer, Harald Schennach, Judith Loeffler-Ragg, Rosa Bellmann-Weiler, Ivan Tancevski, Günter Weiss, Markus Anliker, Andrea Griesmacher, Gregor Hoermann

## Abstract

**Objectives:** Serological tests detect antibodies against Severe Acute Respiratory Syndrome Coronavirus 2 (SARS-CoV-2) in the ongoing coronavirus disease-19 (COVID-19) pandemic. Independent external clinical validation of performance characteristics is of paramount importance.

**Methods:** Four fully automated assays, Roche Elecsys Anti-SARS-CoV-2, Abbott SARS-CoV-2 IgG, Siemens SARS-CoV-2 total (COV2T) and SARS-CoV-2 IgG (COV2G) were evaluated using 350 pre-pandemic samples and 700 samples from 245 COVID-19 patients (158 hospitalized, 87 outpatients).

**Results:** All tests showed very high diagnostic specificity. Sensitivities in samples collected at least 14 days after disease onset were slightly lower than manufacturers’ claims for Roche (93.04%), Abbott (90.83%), and Siemens COV2T (90.26%), and distinctly lower for Siemens COV2G (78.76%). Concordantly negative results were enriched for immunocompromised patients. ROC curve analyses suggest a lowering of the cut-off index for the Siemens COV2G assay. Finally, the combination of two anti-SARS-CoV-2 antibody assays is feasible when considering borderline reactive results.

**Conclusions:** Thorough on-site evaluation of commercially available serologic tests for detection of antibodies against SARS-CoV-2 remains imperative for laboratories. The potentially impaired sensitivity of the Siemens COV2G necessitates a switch to the company’s newly filed SARS-CoV-2 IgG assay (sCOVG) for follow-up studies. A combination of tests could be considered in clinical practice.

## Introduction

Coronavirus disease 2019 (COVID-19) is caused by the Severe Acute Respiratory Syndrome Virus 2 (SARS-CoV-2) (1, 2), and was first described in China in December 2019 and declared pandemic by the WHO on March 11, 2020 (3). As of November 24, 2020 more than 59 million confirmed cases and almost 1.4 million deaths have been reported worldwide (for Austria: more than 250,000 confirmed cases and almost 2,500 deaths) (4). The total prevalence is estimated to be higher due to unrecognized infections (5). The gold standard for the primary diagnosis of acute SARS-CoV-2 infection remains the specific detection of viral RNA by molecular methods, including reverse transcription polymerase chain reaction (RT-PCR) (6, 7). However, molecular methods have some limitations, in particular the relatively short time frame of detectability post symptom onset and thus possible false negative results in patients who present with low viral load at later stages of the disease course (8-10). In contrast, the presence of SARS-CoV-2 antibodies indicates a recent or prior exposure to the virus (11). Serological tests are therefore important to assess the seroprevalence for monitoring the epidemiology of SARS-CoV-2 infection at the population-level. In view of a still relatively low seroprevalence in many countries, specificities of the assays are crucial for epidemiologic studies and the positive predictive value of a result (12). Furthermore, serologic testing in addition to PCR may aid to increase the accuracy of diagnosis, in particular when seroconversion is documented in consecutive blood samples (8, 13). Moreover, SARS-CoV-2 antibody tests are used to select eligible donors for convalescent plasma (14). Finally, serologic tests are used in SARS-CoV-2 vaccine studies to estimate the immunological response to vaccination (15). The durability of the antibody response as well as the extent and duration of immunity against reinfection with SARS-CoV-2 are still under investigation (16).

Immune response to SARS-CoV-2 includes cell-mediated (17) and antibody-mediated immunity (13). The SARS-CoV-2 genome encodes four major structural proteins, the spike (S), envelope (E), membrane (M) and nucleocapsid (N) proteins, and several accessory proteins (18, 19). The S protein with its receptor-binding domain (RBD) and the N protein are widely used as antigens to assess the humoral immune responses by detection of specific antibodies (20-22). Serological assays currently used include neutralizing antibody assays, enzyme-linked immunosorbent assays (ELISA), automated chemiluminescence immunoassays (CLIA) and lateral flow immunoassays (LFIA) (21, 23). Distinct serologic assays detect different antibody isotypes: IgG, IgM, IgA or all isotypes simultaneously (“total antibodies”). ELISA and CLIA raw data are often calculated as index (in relation to a known sample such as provided control material) and reported as positive or negative depending on a predefined cut-off value.

A rapidly growing number of commercial SARS-CoV-2 antibody assays is available. Whereas the manufacturer of an *in vitro* diagnostic (IVD) device is obliged to declare performance specifications of the test, the need for independent validation of commercial assays in clinical settings has been highlighted in systematic reviews of the literature (24, 25). On the one hand, medical laboratories have to verify that they meet the performance specifications. On the other hand, small sample sizes and a lack of samples from patients with mild to moderate clinical course represent a potential bias in performance studies (26, 27). Our study aimed to evaluate four SARS-CoV-2 antibody assays on three fully automated large-scale laboratory analyzers manufactured by Abbott, Roche, and Siemens, respectively. To or knowledge this is the first published external validation of the Siemens SARS-CoV-2 IgG (COV2G) antibody test.

## Materials and methods

### Patients and study design

The present study was performed at the Central Institute of Clinical and Chemical Laboratory Diagnostics at the University Hospital of Innsbruck as part of the clinical evaluation of different SARS-CoV-2 serologic assays. All procedures performed in the present study involving human participants were in accordance with the ethical standards of the Institutional and/or National Research Committee and with the 1964 Helsinki declaration and its later amendments and were approved by the ethics committee of the Medical University of Innsbruck (ethics commission numbers: 1103/2020, 1167/2020).

245 patients with RT-PCR confirmed SARS-CoV-2 infection were included: hospitalized COVID-19 patients at the University Hospital of Innsbruck, reconvalescent COVID-19 patients with persistent cardio-pulmonary damage participating in a prospective observational study (CovILD-study, ClinicalTrials.gov number, NCT04416100) and reconvalescent persons volunteering as plasma donors at the Central Institute for Blood Transfusion and Immunology. According to the clinical course, patients were grouped as outpatients, hospitalized patients at the general ward, or hospitalized patients at the intensive care unit (ICU) ward. The patients’ characteristics are shown in Table 1. In total, 700 patient samples were assessed. 75 patients had one, 66 patients two, 34 patients three, 23 patients four, 24 patients five and 23 patients six or more blood draws. Disease onset was defined as onset of clinical symptoms compatible with COVID-19 infection. Symptom onset was determined by a questionnaire in convalescent donors and by reviewing individual health records in the other patients. If the patient was asymptomatic or the date of symptom onset was not available (n = 35 patients, 15.2%; corresponding to 81 samples, 11.6%), the date of the first positive SARS-CoV-2 RT-PCR was used instead. The median time span between symptom onset and RT-PCR-based diagnosis was 5 days.

**Table 1.**
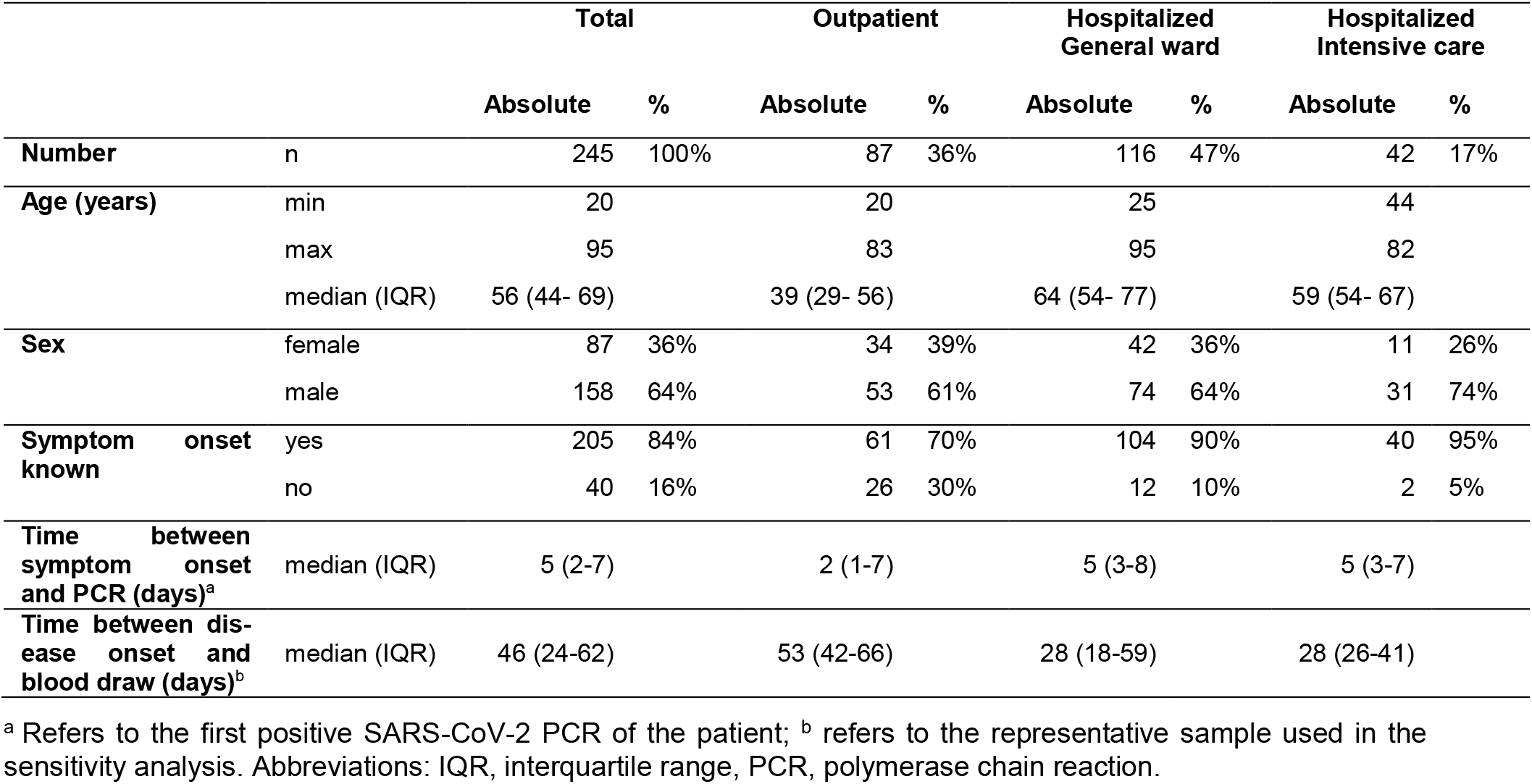
Characteristics of COVID-19 patients

|  |  | Total |  | Outpatient |  | Hospitalized<br>General ward |  | Hospitalized<br>Intensive care |  |
| --- | --- | --- | --- | --- | --- | --- | --- | --- | --- |
|  |  | Absolute | % | Absolute | % | Absolute | % | Absolute | % |
| **Number** | n | 245 | 100% | 87 | 36% | 116 | 47% | 42 | 17% |
| **Age (years)** | min | 20 |  | 20 |  | 25 |  | 44 |  |
|  | max | 95 |  | 83 |  | 95 |  | 82 |  |
|  | median (IQR) | 56 (44- 69) |  | 39 (29- 56) |  | 64 (54- 77) |  | 59 (54- 67) |  |
| **Sex** | female | 87 | 36% | 34 | 39% | 42 | 36% | 11 | 26% |
|  | male | 158 | 64% | 53 | 61% | 74 | 64% | 31 | 74% |
| **Symptom onset known** | yes | 205 | 84% | 61 | 70% | 104 | 90% | 40 | 95% |
|  | no | 40 | 16% | 26 | 30% | 12 | 10% | 2 | 5% |
| **Time between symptom onset and PCR (days)<sup>a</sup>** | median (IQR) | 5 (2-7) |  | 2 (1-7) |  | 5 (3-8) |  | 5 (3-7) |  |
| **Time between disease onset and blood draw (days)<sup>b</sup>** | median (IQR) | 46 (24-62) |  | 53 (42-66) |  | 28 (18-59) |  | 28 (26-41) |  |

<sup>a</sup> Refers to the first positive SARS-CoV-2 PCR of the patient; <sup>b</sup> refers to the representative sample used in the sensitivity analysis. Abbreviations: IQR, interquartile range, PCR, polymerase chain reaction.

In addition, 350 archived samples drawn in the pre-COVID-19 era were used to validate the specificity of the assays. In detail, 274 unselected samples dated from February 2017 to November 2019, 51 samples from hospitalized patients with bacterial pneumonia and 25 samples from patients with rheumatologic diseases (14 rheumatoid arthritis, six spondylarthritis, three connective tissue disease, two late onset rheumatoid arthritis) drawn before the COVID-19 era were examined. Additionally, an intravenous immunoglobulin formulation (Privigen®, 100 mg/ml, CSL Behring AG, Bern, Switzerland) composed of pre-pandemic pooled immunoglobulins (mainly IgG) of a large number of healthy donors from the US, which should by definition yield negative SARS-CoV-2 antibody results, was used for antibody assay evaluations.

### Sample preparation

Blood samples were drawn according to routine clinical procedures. Upon centrifugation, serum specimens for antibody determination were kept at 4°C if analyses were conducted within 7 days or stored at −20°C in the case analyses were conducted at a later time point. Frozen samples were thawed and centrifuged prior to antibody determination.

### Anti-SARS-CoV-2 assays

We evaluated the performance of the following fully automated CLIA tests on high throughput random access analyzers widely available in medical laboratories: Roche Elecsys Anti-SARS-CoV-2 assay on the Cobas e602 platform (Roche Diagnostics, Rotkreuz, Switzerland), Abbott SARS-CoV-2 IgG assay on the Architect i2000SR platform (Abbott Laboratories Abbott Park, IL, USA), Siemens SARS-CoV-2 total (COV2T) and SARS-CoV-2 IgG (COV2G) on the Advia Centaur XP platform (Siemens, Munich, Germany). All samples were processed according to the manufacturers’ procedures with the specified controls and calibrators by trained laboratory staff. Test characteristics given in the manufacturers’ product information are summarized in Supplemental Table S1.

### Data analysis and statistics

Specificity was analyzed on 350 archived samples drawn in the pre-COVID-19 era and sensitivity on samples from patients with RT-PCR confirmed SARS-CoV-2 infection. Only one sample per patient was subjected to sensitivity analysis to avoid bias due to multiple testing. Patients were included if at least one sample dating between day 14 and day 120 after disease onset was available. In case of multiple samples available per patient, the sample closest to day 28 was included in the sensitivity analysis.

Statistical analyses were performed using MedCalc, version 11.5.1.0 (MedCalc Ltd., Ostend, Belgium) and Excel 2016 (Microsoft, Redmont, USA). Median and interquartile range (IQR) were used as descriptive measures of metric data. Categorical data are given as counts and percentages. 95% confidence intervals (CI) for proportions were calculated according to the Clopper-Pearson exact method. The difference between categorical data was assessed using Chi-square test (McNemar’s test for paired data, “N-1” Chi-squared test for unpaired proportions). Receiver operating characteristic (ROC) curve analysis was performed using the DeLong method. The concordance correlation coefficient was calculated according Lin. Statistical significance was defined at a level of 0.05.

## Results

### Specificity

Of the 350 pre-pandemic samples included, 341 were analyzed with the Roche Elecsys Anti-SARS-CoV-2 assay, 298 with Abbott SARS-CoV-2 IgG, 288 with Siemens SARS-CoV-2 total (COV2T), and 191 with Siemens SARS-CoV-2 IgG (COV2G). Using the manufacturers’ cut-offs, the specificity ranged from 99.33% to 100.00% (Roche 99.71%, Abbott 99.33%, Siemens COV2T 99.65%, Siemens COV2G 100.00%; Table 2, Supplemental Table S2). The small number of false positive samples (Supplemental Table S3) showed no overlap between the tests as none of the samples was tested positive in more than one assay. No false positive result was observed in the sub-cohorts of pre-pandemic samples from patients with bacterial pneumonia (n=51) or rheumatologic diseases (n=25). Borderline cross reactivity when measuring undiluted intravenous immunoglobulin formulation (Privigen®) was found only for Siemens SARS-CoV-2 IgG (Index: 1.09). However when diluting Privigen® 1:50 with SARS-CoV-2 negative serum or sodium chloride 0.9% (reflecting a more physiological immunoglobulin concentration) all assays shielded negative results (Supplemental Table S4).

**Table 2.** Sensitivity and specificity of the assays

|  |  | Investigated patient cohort | Manufacturers' claims |
| --- | --- | --- | --- |
| **Roche Elecsys Anti-SARS-CoV-2** | Sensitivity (n=230) | 93.04% (88.95-95.97)<sup>a</sup> | 99.50% (97.00-100.00)<sup>b</sup> |
|  | Specificity (n=341) | 99.71% (98.36-99.95) | 99.80% (99.69-99.88) |
| **Abbott SARS-CoV-2 IgG** | Sensitivity (n=228) | 90.79% (86.33-93.90)<sup>a</sup> | 100.0% (95.89-100.00)<sup>c</sup> |
|  | Specificity (n=298) | 99.33% (97.59-99.82) | 99.63% (98.98-99.89) |
| **Siemens SARS-CoV-2 total (COV2T)** | Sensitivity (n=195) | 90.26% (85.28-93.67)<sup>a</sup> | 100.0% (92.45-100.00)<sup>b</sup> |
|  | Specificity (n=288) | 99.65% (98.06-99.94) | 99.81% (99.41-99.96) |
| **Siemens SARS-CoV-2 IgG (COV2G)** | Sensitivity (n=193) | 78.76% (72.45-83.94)<sup>a</sup> | 100.00% (91.59-100.00)<sup>b</sup> |
|  | Specificity (n=191) | 100.00% (98.03-100.00) | 99.89% (99.61-99.99) |

Sensitivity in the investigated cohort was evaluated using samples of SARS-CoV-2 PCR-confirmed patients dating $\geq 14$ days after disease onset and the sample closest to day 28 after disease onset was chosen (one sample per patient). Specificity was determined on pre-pandemic samples. 95% confidence intervals are shown in brackets. <sup>a</sup>: ≥ day 14 after disease onset; <sup>b</sup>: ≥ day 14 after first PCR-positivity; <sup>c</sup>: ≥ day 14 after symptom onset

### Sensitivity

From 230 (93.9%) of the 245 PCR-confirmed COVID-19 patients, a sample dating between day 14 and day 120 after disease onset was available for analysis. The median time between disease onset and blood sampling was 46 days (IQR 24-62, range 14-120; Table 1, Supplemental Figure S1). All single results of the 230 samples are shown in Supplemental Table S5. Using the manufacturers’ cut-offs, the observed sensitivity of the assays ranged from 78.76% to 93.04% (Roche Elecsys Anti-SARS-CoV-2 93.04%, Abbott SARS-CoV-2 IgG 90.79%, Siemens SARS-CoV-2 total 90.26%, Siemens SARS-CoV-2 IgG 78.76%; Table 2, Supplemental Table S6). Compared to the COV2G assay, the Roche (p<0.0001), Abbott (p=0.0001) and COV2T (p<0.0001) tests were significantly more sensitive.

In an exploratory analysis, we assessed the sensitivity of the assays stratified for gender, age, and disease severity (Supplemental Table S6). When stratifying for gender, sensitivity was higher for males than for females within the Abbott (male: 94.52%, female: 84.15%; p=0.0095) and Siemens COV2G (male: 87.30%, female: 62.69%; p=0.0001) assays (Figure 1A). Stratified for patient age, sensitivity for Siemens COV2G was lower in the age group 18-49 years (63.93%) compared to patients aged 50-69 years (86.32%) and 70-100 years (83.78%) (Figure 1B). Next, we stratified the patients according to intensity of care needed (outpatients, n=85; hospitalized patients in general ward, n=107; and ICU patients, n=39) as a surrogate for disease severity. For outpatients sensitivity ranged from 64.86% to 91.76% (Roche 91.76%, Abbott 85.88%, Siemens COV2T 88.89%, Siemens COV2G 64.86%). For hospitalized patients at the general ward sensitivity ranged from 84.27% to 93.40% (Roche 93.40%, Abbott 93.33%, Siemens COV2T 90.00%, Siemens COV2G 84.27%). For ICU patients sensitivity ranged from 93.94% to 96.67% (Roche 94.87%, Abbott 94.74%, Siemens COV2T 93.94%, Siemens COV2G 96.67%). In particular, the COV2G test had a significantly lower sensitivity in outpatient compared to general ward or ICU patients (Figure 1C).

**Figure 1.**
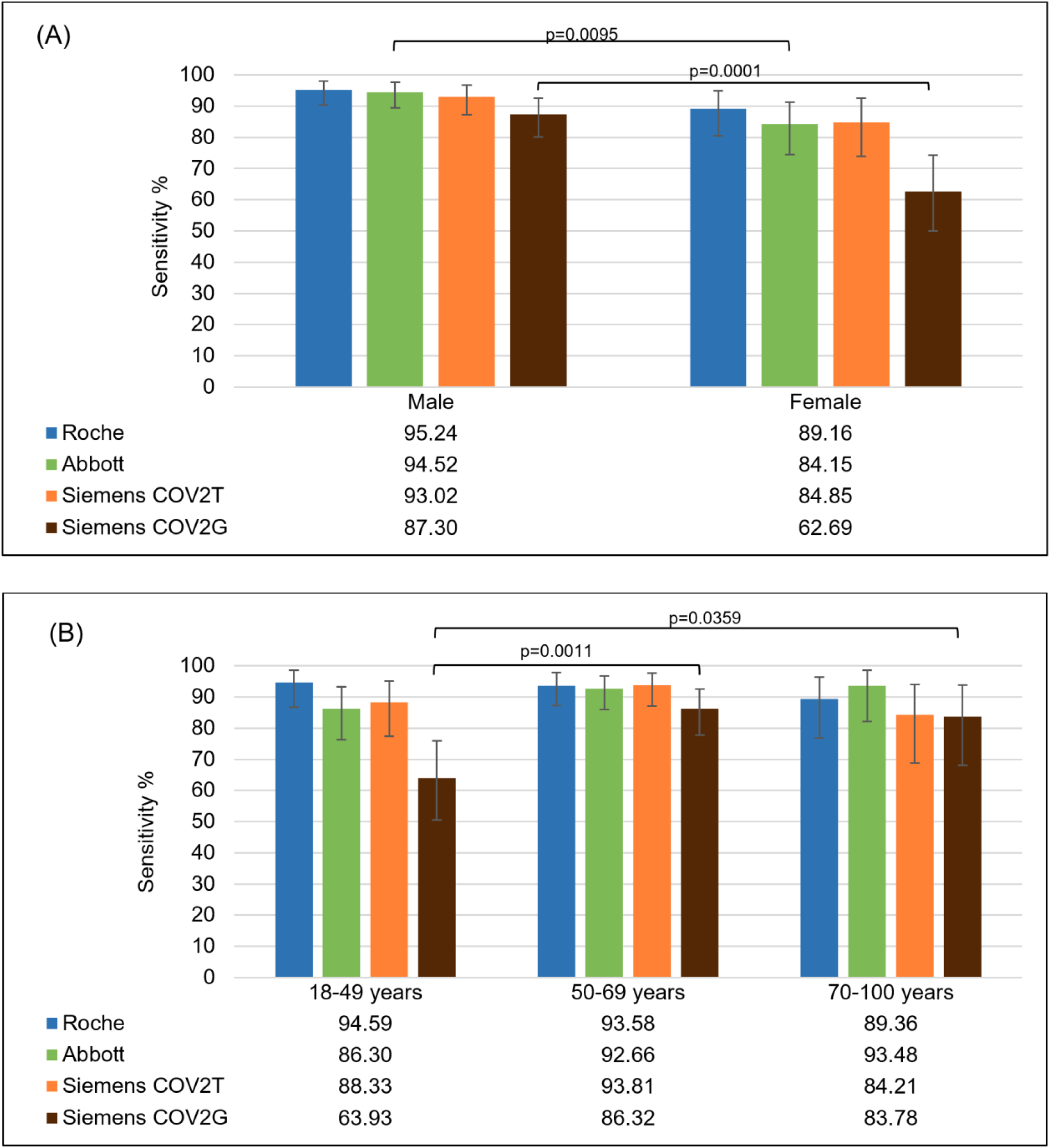
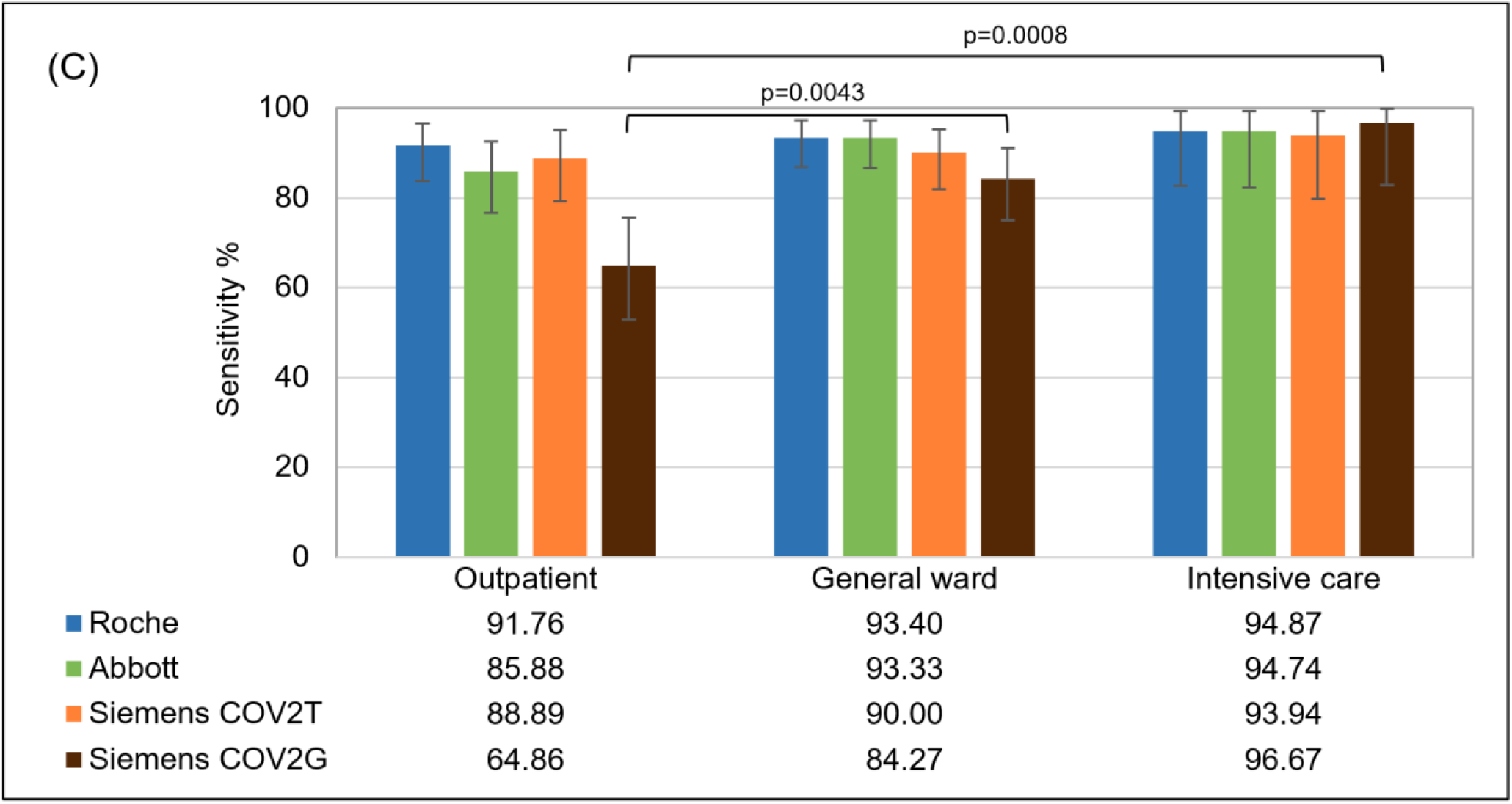
Subgroup analyses of sensitivity according to gender, age and severity of disease. Comparison of the sensitivity of the four investigated tests in representative samples of 230 patients with PCR-confirmed COVID-19: Roche (Roche Elecsys Anti-SARS-CoV-2, blue), Abbott (Abbott SARS-CoV-2 IgG, green), Siemens COV2T (Siemens SARS-CoV-2 total, orange) and Siemens COVG (Siemens SARS-CoV-2 IgG, brown). Results were analyzed stratified for gender (A), age (B) and severity of disease (C).

To address potential reasons for unexpected negative results, clinical records of patients tested negative with the majority of SARS-CoV-2 antibody tests were analyzed in detail. Out of the 230 patient samples included in the primary endpoint analysis, 13 samples gave negative results in all performed tests and two samples in all but one assay (one was positive only in the Abbott and one only in the Siemens COV2T test). Seven out of those 15 patients were under immunosuppressive treatment not related to COVID-19. Of those, three were under ongoing chemotherapy due to malignancy, two patients had ongoing anti-CD20 therapy with obinutuzumab or rituximab due to lymphoma, one patient received methotrexate due to rheumatoid arthritis and another patient was under treatment with corticosteroids and rituximab because of myasthenia gravis. The other eight patients did not have relevant immunosuppressive conditions or therapies and in all those patients total serum IgG, IgM and IgA immunoglobulins were within the normal range.

### Positivity rate across the course of disease

To estimate the time to positivity after disease onset, we analyzed all 700 samples from 245 PCR-confirmed COVID-19 patients and stratified the positivity rate for the time from disease onset (groups days 0-6, 7-13, 14-20, 21-40 and >40). For all tests, the positivity rate increased from week one (range 19.51% to 32.14%) to week two (range 40.62% to 54.64%) and week three (range 80.85% to 95.33%). While the positivity rate after week three slightly decreased for Roche and Abbott, the positivity rate of samples for Siemens COV2T and Siemens COV2G increased from day 14-20 to day 21-40 and dropped again after day 40 (Figure 2, Supplemental Table S7, Supplemental Figure S2). The latter effect might be confounded by the higher number of out-patient samples in the group of samples obtained >40 days from disease onset (Supplemental Figure S1).

**Figure 2.**
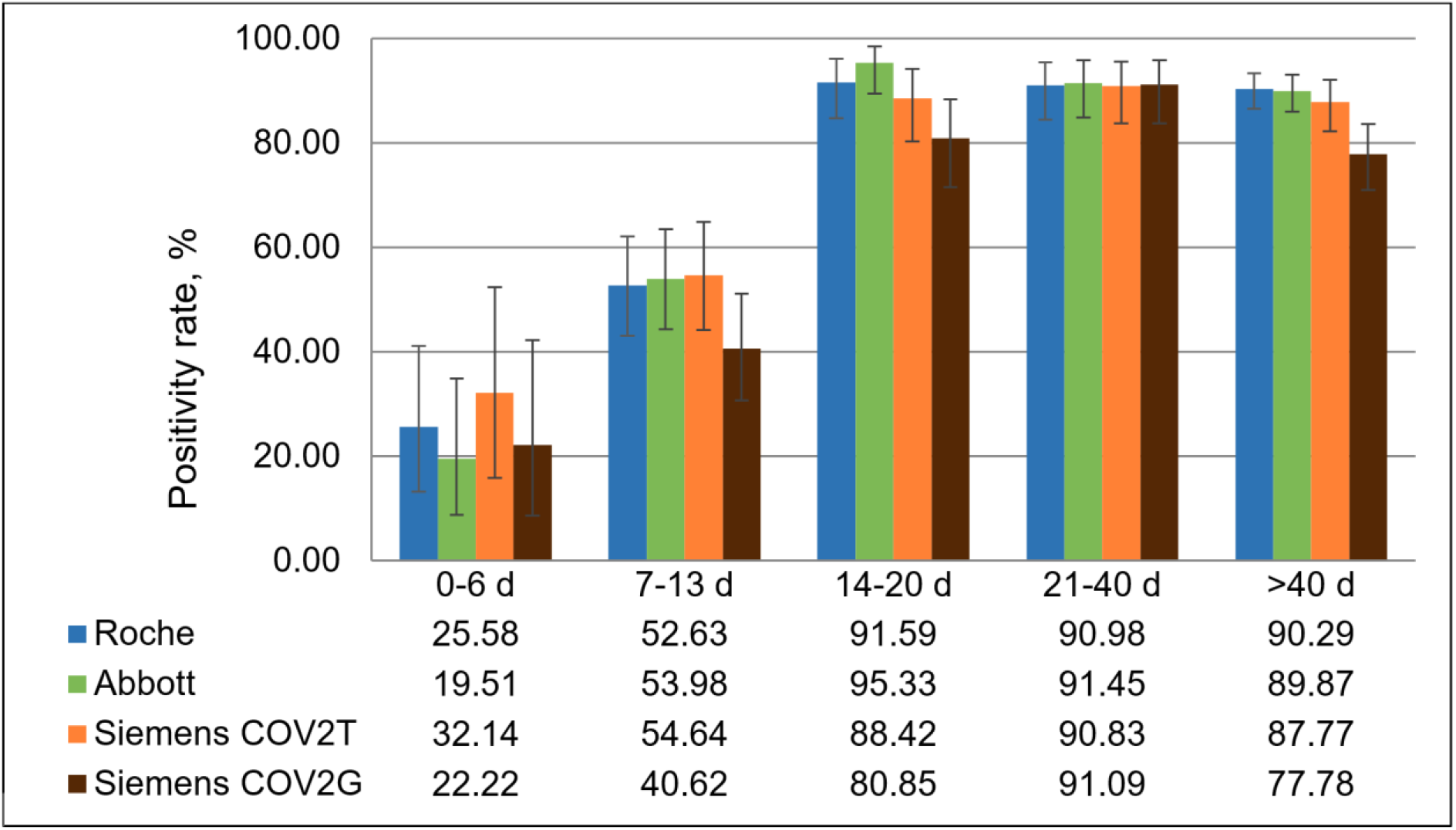
Positivity rate across the course of disease. All 700 samples from 245 PCR-confirmed SARS-CoV-2 infected patients were subjected to antibody determination with Roche (Roche Elecsys Anti-SARS-CoV-2, n=695, blue), Abbott (Abbott SARS-CoV-2 IgG, n=684, green), Siemens COV2T (Siemens SARS-CoV-2 total, n=517, orange) and Siemens COVG (Siemens SARS-CoV-2 IgG, n=498, brown). The positivity rate of the respective tests is shown in dependence of the time between disease onset and blood draw in days (d).

### ROC curve analysis

ROC curve analysis revealed area under the curve (AUC) values of 0.984 for Roche Elecsys Anti-SARS-CoV-2, 0.982 for Abbott SARS-CoV-2 IgG, 0.975 for Siemens SARS-CoV-2 total (COV2T) and 0.966 for Siemens SARS-CoV-2 IgG (COV2G) (Figure 3, Supplemental Table S8). When comparing the ROC curves, the AUC of the COV2G test was significantly smaller than the AUC of Roche (p=0.0151) and Abbott (p=0.0174).

**Figure 3.**
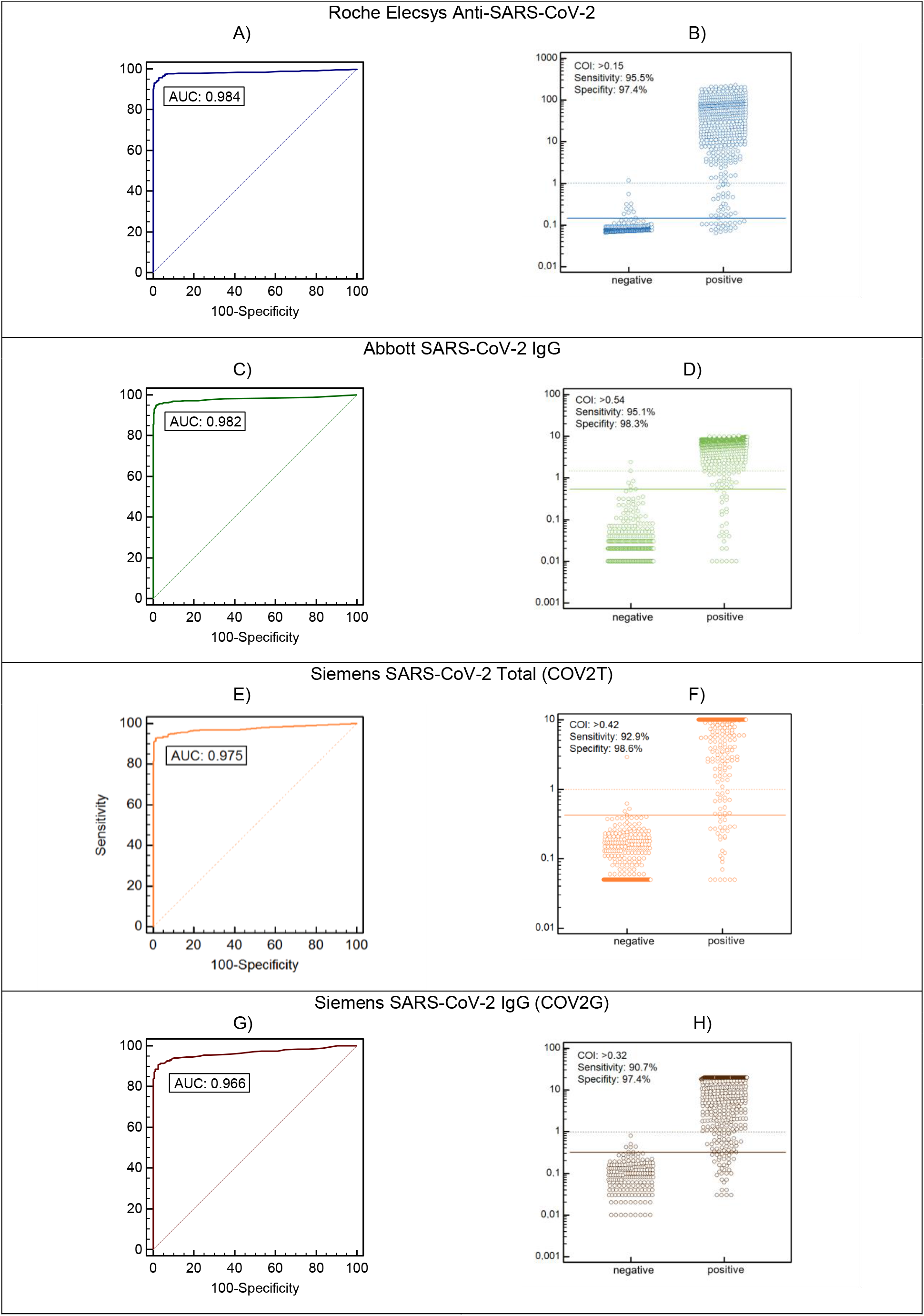
ROC curve analysis and modified cut-off indexes. ROC curves (left) and corresponding raw data indexes (right) for the investigated tests from Roche (Roche Elecsys Anti-SARS-CoV-2, A-B, blue), Abbott (Abbott SARS-CoV-2 IgG, C-D, green), Siemens COV2T (Siemens SARS-CoV-2 total, E-F, orange) and Siemens COV2G (Siemens SARS-CoV-2 IgG, G-H, brown). Analyses are based on pre-pandemic (=negative) samples (samples, Roche: 341, Abbott: 298, Siemens COV2T: 288, Siemens COV2G: 191) and all the samples of SARS-CoV-2 PCR-confirmed patients dating ≥14 days after disease onset (samples, Roche: 537, Abbott: 530, Siemens COV2T: 392, Siemens COV2G: 375). The horizontal lines in the dot plots represent the manufacturers’ cut off index (dashed line, Roche and Siemens: 1.0, Abbot: 1.4) and the modified cut off index according to ROC curve analysis (solid line).

In an exploratory analysis, we asked if the assay cut-offs could be modified to improve the sensitivity. The proposed cut-off indexes (COI) based on the maximum sum of sensitivity and specificity in ROC curve analysis were below the manufacturers’ COI: >0.15 for Roche, >0.54 for Abbott, >0.42 for Siemens COV2T and >0.32 for Siemens COV2G (Figure 3). Using these optimized COI instead of the manufacturers’ COI, sensitivity improved from 93.04% to 95.65% for Roche, from 90.79% to 95.18% for Abbott, from 90.26% to 92.31% for Siemens COV2T and from 78.76% to 89.64% for Siemens COV2G. However, specificity of the single assays diminished from 99.71% to 97.36% for Roche, from 99.33% to 98.32% for Abbott, from 99.65% to 98.61% for Siemens COV2T and from 100.00% to 97.38% for Siemens COV2G when these modified COI were applied (Supplemental Table S9).

### Concordance between results of different antibody assays indicates a potential for test combinations

When considering all 1,050 samples the concordance correlation coefficient between the four assays ranged from 77.45% to 92.49% (Supplemental Table S10). Of the 350 pre-pandemic samples included in our study, 235 were tested with all three assays. 233 of these samples (99.1%) were tested negative with all three assays. In two (0.9%) samples, only the Abbott test yielded a positive result. To compare the sensitivity of the assays in detail, we restricted the analysis to a single sample per patient obtained between day 14 and day 120 after disease onset as described above. 189 samples of PCR-confirmed COVID-19 patients were tested with the four assays Roche, Abbott, and Siemens (COV2T and COV2G). 145 (76.7%) of them were positive with all four assays, and 11 (5.8%) were found negative in all four assays. Of note, 18 samples (9.5%) were negative only with COV2G (Figure 4). Due to the unexpected low sensitivity of the Siemens COV2G test, we restricted further analyses on combinatorial approaches to the three assays Roche Elecsys Anti-SARS-CoV-2, Abbott SARS-CoV-2 IgG and Siemens SARS-CoV-2 total (COV2T).

**Figure 4.**
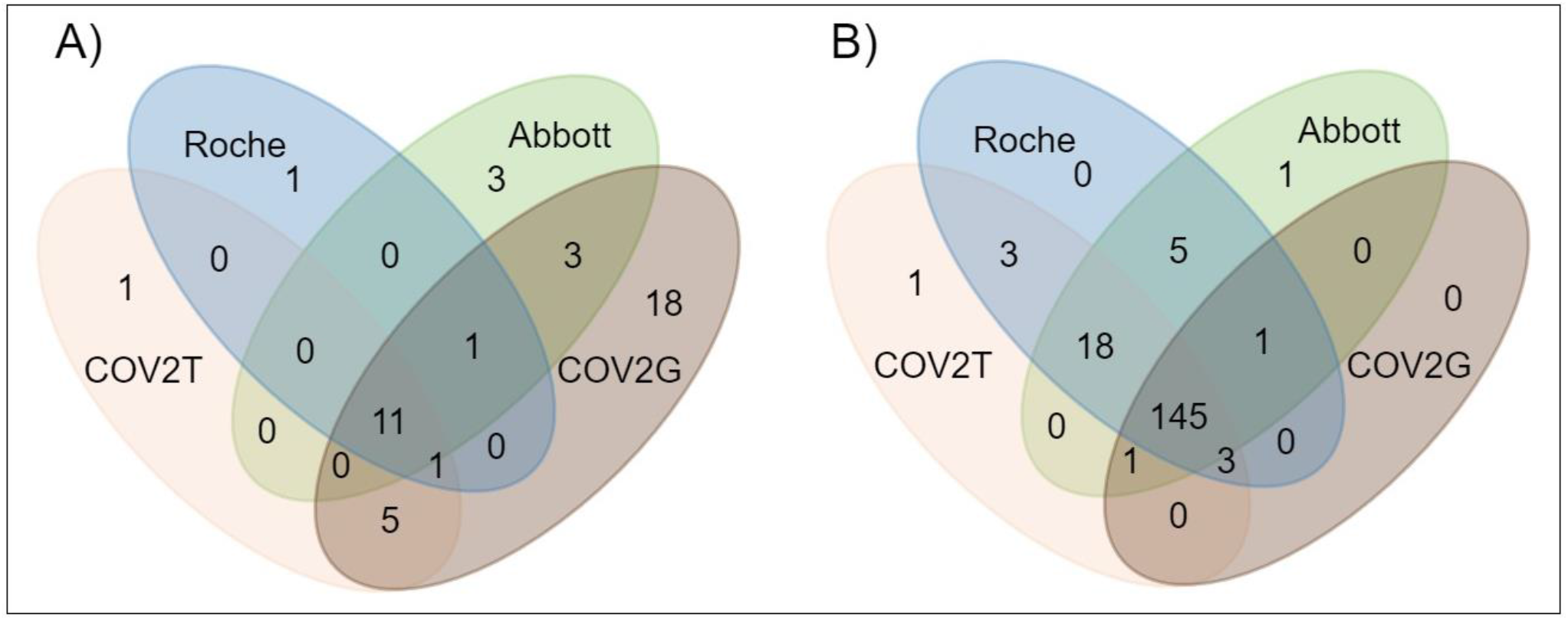
Concordance of the SARS-CoV-2 antibody results in PCR-confirmed patients. Overlap of negative (A) and positive (B) antibody test results of different assays in SARS-CoV-2 PCR-confirmed COVID-19 patients (samples dating ≥14 days after disease onset; one representative sample per patient; a total of 189 samples in which data from all four assays were available). Abbreviations: Abbott, Abbott SARS-CoV-2 IgG; Roche, Roche Elecsys Anti-SARS-CoV-2; COV2G, Siemens SARS-CoV-2 IgG; COV2T, Siemens SARS-CoV-2 total.

Finally, we asked whether a combination of two antibody tests could be useful to further improve the clinical performance of serologic SARS-CoV-2 tests. When combining any two out of the Roche, Abbott and Siemens COV2T assays, specificity improves to 100.00%, regardless of the combination used. However, sensitivity dropped to <90% when considering only samples with concordantly positive results in both tests (Table 3). When we used the modified COI instead of the manufacturers’ COI, sensitivity of the combinations improved to 95.18% for Roche and Abbott (manufacturers’ COI: 89.91%), 92.31% for Roche and Siemens COV2T (manufacturers’ COI: 89.23%), and 92.27% for Abbott and Siemens COV2T (manufacturers’ COI: 86.60%). In contrast, the specificity of the combined analyses was not affected by the lowering of the COI (Table 3).

**Table 3.** Sensitivity and specificity for the combination of two tests

|  |  | **Manufacturers' COI** | **Modified COI** |
| --- | --- | --- | --- |
| **Roche + Abbott** | Combined sensitivity (n=228) | 89.91 (85.32-93.18%) | 95.18% (91.57-97.28%) |
|  | Combined specificity (n=291) | 100.00% (98.70-100.00%) | 100.00% (98.70-100.00%) |
| **Roche + Siemens COV2T** | Combined sensitivity (n=195) | 89.23% (84.10-92.85%) | 92.31% (87.70-95.28%) |
|  | Combined specificity (n=282) | 100.00% (98.66-100.00%) | 100.00% (98.66-100.00%) |
| **Abbott + Siemens COV2T** | Combined sensitivity (n=194) | 86.60% (81.09-90.69%) | 92.27% (87.64-95.26%) |
|  | Combined specificity (n=241) | 100.00% (98.43-100.00%) | 100.00% (98.43-100.00%) |

Sensitivity and specificity for the combination of assays (double positive samples were regarded as positive, single or double negative samples were regarded as negative) using different cut-off indexes (COI): the COI according to the manufacturer and a modified COI according to ROC curve analysis of the results of our cohort. Sensitivity in the investigated cohort was evaluated using samples of SARS-CoV-2 PCR-confirmed patients dating $\geq 14$ days after disease onset and the sample closest to day 28 after disease onset was chosen (one sample per patient). Specificity was determined on pre-pandemic samples. 95% confidence intervals are shown in brackets. Abbreviations: Roche, Roche Elecsys Anti-SARS-CoV-2; Abbott, Abbott SARS-CoV-2 IgG; Siemens COV2T, Siemens SARS-CoV-2 total.

## Discussion

In the present study we examined the performance characteristics of four fully automated SARS-CoV-2 antibody CLIA assays focusing on high throughput random access analyzers widely available in many medical laboratories. While the specificity of all assays was well comparable to the performance characteristics provided by the manufacturers, we observed a markedly lower sensitivity in our cohort (78.76% to 93.04% compared to >99%). Similar results have been found in other studies for the Roche, Abbott and the Siemens COV2T assays (28, 29). These findings emphasize the importance of real life data and different clinical scenarios in independent assay validations. While the more subtle differences between observed and expected sensitivity rates for Roche, Abbott and Siemens COV2T were coherent within our study and might be explained by specific characteristics of our patients cohort, the sensitivity of Siemens COV2G was markedly lower in our hands with 78.76% in the total cohort and 64.86% in outpatients. To the best of our knowledge, our study is the first public-available independent evaluation of the COV2G assay and warrants further studies to verify and potentially improve the performance of this test. ROC curve analysis of our results of the Siemens COV2G assay suggests that the cut-off might be too high. Indeed, a number of false negative samples showed borderline reactivity that did not exceed the manufactures’ assay cut-off of 1.0. Lowering the cut-off to 0.32 would improve the sensitivity from 78.76% to 89.64% with modest effect on the assay specificity (100.00% to 97.38%). Of course, this post-hoc change of the cut-off would require to repeat the evaluation of sensitivity and specificity for the Siemens COV2G assay in a large cohort. Indeed, the company has just recently filed an FDA-application for approval of a new SARS-CoV-2 IgG assay (sCOVG) with improved sensitivity. Our results indicate that potential differences in the assay performance between the Siemens COV2G and the new sCOVG assays need to be considered when evaluating sequential samples of patients.

Interestingly, we found lower sensitivities in female vs. male patients with all four assays, whereby a statistical significant difference was found only in the Abbott and Siemens COV2G assays. In a sub-analysis this trend for a gender difference was found in all three different courses of disease (outpatient, general ward, ICU; data not shown). This is grossly in line with the findings of Korte et al., which also showed that men produce higher amounts of anti SARS-CoV-2 IgG and IgA after SARS-CoV-2 infection (30). This finding should also be included in the interpretation of single antibody test results.

Inconsistent results have been reported regarding the association between antibody titers and disease severity. For example in a serosurvey in health care workers of the Veneto Region of Italy, Plebani et al. found that symptomatic individuals were 100% SARS-CoV-2 antibody positive, whilst only in 58% of asymptomatic carriers antibodies were detectable (31). Phipps et al. could not find an association of IgG (Abbott) and IgM (in-house) antibody response and disease severity, however, patients were segregated in ICU vs. non-ICU care and unlike to the study of Plebani and our study no asymptomatic or outpatient populations, respectively, were described separately (32). The importance of including patients with mild disease course in antibody evaluation studies has already been highlighted (26). We did not observe a major difference between the positivity rates for outpatients, hospitalized patients and ICU patients for the Roche, Abbott and the Siemens COV2T assays. In contrast, the sensitivity rates for the Siemens COV2G were significantly lower in outpatients compared to hospitalized patients. The stringent cut-off of this particular assay might affect the results in patients with low antibody levels. However, none of the assays evaluated was optimized for quantification of antibody titers and quantitative analysis was limited by the dynamic range of the tests. Thus, our study was not designed to assess quantitative differences in antibody levels between patients.

Notably, absence of reactivity in serological assays could either reflect antibody test performance or the biological absence of antibodies in individuals as no clear gold standard for the evaluation of SARS-CoV-2 antibody tests exists (33). As samples from 13 patients (5.7%) were constantly negative in all assays, we conclude that the biological absence of antibodies was a relevant factor in our cohort. The biological absence of antibodies in individuals with RT-PCR confirmed SARS-CoV-2 infection might be explained by immediate infection clearance in the naso-pharyngeal space as a consequence of low viral exposure and/or effective immune function which does not induce a systemic immune response but results in detection of viral RNA by RT-PCR. While no data on T-cell mediated immune response were available in our patients, detailed clinical meta-data allowed to correlate the absence of humoral immune response with immunosuppression. Indeed, 5 of those 13 patients with constantly negative antibody tests were found being immunocompromised what likely explains the absence of a detectable systemic humoral immune response. The proportion of immunocompromised patients seems to affect the overall sensitivity rate in our cohort, and our findings warrant for careful interpretation of SARS-CoV-2 antibody results particularly in those.

SARS-CoV-2 seroprevalence is still used as a measure to estimate the true number of affected people during the pandemic. Such studies found seropositivity for SARS-CoV-2 antibodies in 2.4% (n=61,437) of residents in Wuhan in mid-May (34), 0.9% (n=3,186) in German regular blood donors from March to June (35), 1.0% (n=2,500) in Greece university personnel and students during June-July (36), and 4.6% (n=5,933) of health care workers till end of May at the University-Hospitals of Padova and Verona, Italy (31). Thus, a very high diagnostic specificity of serologic tests is crucial to minimize false positive results and improve the positive predictive value (PPV). In line with the performance characteristics provided by the manufacturers, we observed a false positive rate <1% for all tests using samples from the pre-pandemic era. However, even a small false positive rate substantially effects the PPV in a low-prevalence situation (12). It has thus been suggested to combine the results of two different SARS-CoV-2 antibody tests to further improve specificity and PPV of serologic testing (37, 38). As a prerequisite for this strategy, it has been shown that the vast majority of false positive results occurred independently with singular anti-SARS-CoV-2 CLIA assays whereas systematic false positive samples affecting multiple assays are very rarely observed (12). This is also in line with our observation. Although the number of false positive samples in our study is rather small, we observed not a single sample being false positive in more than one CLIA assay. In contrast, the concordance of false negative samples between the Roche, Abbott and Siemens COV2T assays was rather high and yielded in a combined sensitivity of >86% for any of these combinations. While singular tests have been optimized for maximal specificity, a combinatorial testing approach could allow to lower the thresholds to recognize borderline reactive samples without impairing specificity (38). With the lower cut-offs according to the ROC analysis, the combined sensitivity improved to >92% for any combination. These results suggest, that borderline reactive results slightly below the threshold of the test could benefit from further analyses with additional CLIA or ELISA assays. However, due to the huge impact of the disease prevalence and pretest probability on predictive values of the assay results, the purpose of the test (detection or exclusion of disease, seroprevalence studies) should be defined before adjustment of thresholds or combining different serologic tests in a certain setting (39).

Limitations of our study include differences in the time between disease onset and serological assessment between patients due to the retrospective design of this assay validation study. To avoid potential biases, we constrained our analysis to samples drawn ≥14 days after onset of COVID-19 specific symptoms in SARS-CoV-2 RT-PCR confirmed patients or two weeks after the first positive RT-PCR result in patients for whom no information on symptom onset was available. The median time of 46 days between disease onset and date of the sample used for the sensitivity analysis fits well to the reported plateau of antibody production against SARS-CoV-2 (40). Thus, our study was designed to assess the maximal sensitivity of antibody tests ≥14 days and did not include the early phase of antibody production within the first days of COVID-19. Potential differences between the performances of the evaluated tests in the symptomatic phase of the disease can thus not be excluded. Likewise, the maximum time between disease onset and sampling was 120 days. The duration of antibody production and immunity after a SARS-CoV-2 infection is a major question. For example, in the study of Long et al. 40% of asymptomatic and 13% of symptomatic patients became seronegative in the early convalescent phase (41). Liu et al. found that SARS-CoV-2 antibodies substantially decreased in about 60 days after symptom onset (42). On the other hand, Isho et al. showed that IgG antibodies to SARS-CoV-2 are maintained in the majority of COVID-19 patients for at least three months post symptom onset (43). However, among others, the observed discrepancies may also be due to differences in the serologic assays used in the different studies. In our study, no substantial decrease of seropositivity was found within 120 days after disease onset. Further studies are needed to assess the performance of different serological tests in longitudinal analysis. In this regard, quantitative measurements of antibody titers and neutralization or pseudo-neutralization assays will be useful to monitor the course of the humoral immune response against SARS-CoV-2 in detail. Another limitation of our study was that only adult patients with COVID-19 have been included. For example, Pierce et al. found that serum neutralizing antibody titers were higher in adults compared to pediatric patients with COVID-19 (44). Additional studies are needed to evaluate the performance of the assays tested here for children.

In summary, we independently evaluated the performance of four widely available commercial SARS-CoV-2 antibody assays in an adult COVID-19 cohort including both patients with critical to severe as well as mild courses of the disease. While all assays met the desired specificity criteria, we observed a substantially lower sensitivity compared to the performance reported by the manufacturers. Our study emphasizes the importance of achieving additional performance data in real life including specific populations like immunocompromised patients and asymptomatic carriers. Importantly, our results suggest a limited sensitivity of the Siemens COV2G assay that will be replaced by the newly filed Siemens sCOVG assay. In conclusion, a growing number of fully automated SARS-CoV-2 antibody CLIA assays with sufficient performance characteristics is available for different high throughput analyzers. The selection of specific assays and the interpretation of results should carefully reflect the use case, and a combination of different SARS-CoV-2 antibody assays might be useful.

## Data Availability

See section supplemental data

## Abbreviations

CI: confidence interval
CLIA: chemiluminescence immunoassay
COI: cut-off index
COVID-19: Coronavirus disease 2019
ELISA: enzyme-linked immunosorbent assay
LFIA: lateral flow immunoassay
ICU: intensive care unit
Ig: Immunoglobulin
IQR: interquartile range
ROC: receiver operating characteristic
RT-PCR: reverse transcription polymerase chain reaction

## Acknowledgements

The authors thank Bettina Schatz, Daniel Außerdorfer (both Central Institute of Clinical and Chemical Laboratory Diagnostics, University Hospital Innsbruck) and Gernot Osterer (Siemens) for technical support and are grateful to Manfred Herold (Department of Internal Medicine II, University Hospital Innsbruck) for providing serum samples of patients with rheumatologic diseases.

## Research funding

The study was performed by institutional research funding.

## Author contributions

CI, AE, WP, LL, MA, AG, and GH performed or analyzed serological tests. SS, TS, WM, HS, JL-R, RB-W, IT, and GW obtained clinical data. CI and AE performed statistical analysis. CI, AE, AG, and GH designed the study and wrote the paper. All authors revised and approved the manuscript.

## Competing interests

The authors declare no competing interest.

## Financial

IT was awarded an Investigator-Initiated Study (IIS) grant by Boehringer Ingelheim (IIS 1199-0424).

## Notes

### Competing Interest Statement

The authors have declared no competing interest.

### Funding Statement

no external funding

### Author Declarations

Approved by the ethics committee of the Medical University of Innsbruck (ethics commission numbers: 1103/2020, 1167/2020

### Summary of Updates

1) updated Ref. 12 from "medRxiv" to "Clin Chem. 2020;66(11):1405-13." 2) addition of 1 reference (Ref. #38)

