## Supplemental material for "Evaluation of four commercial, fully automated SARS-CoV-2 antibody tests suggests a revision of the Siemens SARS-CoV-2 IgG assay"

### Supplemental Tables

#### **Supplemental Table S1.** Attributes for the evaluated serologic assays as given by the manufacturers' package inserts.

| Attribute | Elecsys Anti-SARS-CoV-2 | SARS-CoV-2 IgG | SARS-CoV-2 total (COV2T) | SARS-CoV-2 IgG (COV2G) |
|---|---|---|---|---|
| **Manufacturer** | **Roche** | **Abbott** | **Siemens** | **Siemens** |
| **Version of the package insert** | July 2020 | April 2020 | Mai 2020 | June 2020 |
| **CE-IVD status** | Conform | Conform | Conform | Conform |
| **EUA status** | Granted | Granted | Granted | Granted |
| **Method** | ECLIA | CMIA | CLIA | CLIA |
| **Platform** | Cobas e411, e601, e602[a], e801 | ARCHITECT i2000SR [a], i1000SR | ADVIA Centaur XP [a], XPT | ADVIA Centaur XP [a], XPT |
| **Specimen type** | Serum, plasma (Li-heparin, K-EDTA) | Serum, plasma (EDTA) | Serum, plasma (Li-heparin, K-EDTA) | Serum, plasma (Li-heparin, K-EDTA) |
| **Sample volume (without dead volume)** | 20 µl | 25 µl | 50 µl | 10 µl |
| **Detected antibodies** | Total antibodies, including IgG | IgG | Total antibodies, including IgG and IgM | IgG |
| **Solid-phase antigen target** | N protein | N protein | RBD of the S1 protein | RBD of the S1 protein |
| **Result calculation** | Cut-off index | Index (S/Co) | Index | Index |
| **Interpretation** | <1,0: negative<br>≥1,0: positive | <1,4: negative<br>≥1,4: positive | <1,0: negative<br>≥1,0: positive | <1,0: negative<br>≥1,0: positive |
| **Diagnostic specificity*** | 99.80% (99.69-99.88%). n=10,432 | 99.63% (98.98-99.89%). n=1,066 | 99.81% (99.41-99.96%). n=1,589 | 99.89% (99.61-99.99%). n=1,831) |
| **Diagnostic sensitivity*** | *Days after first PCR-positivity*<br>0-6: 60.20% (52.30-67.80%). n=161<br>7-13: 85.30% (78.60-90.60%). n=150<br>≥14: 99.50% (97.00-100.00%). n=185 | *Days after symptom onset*<br><3: 0.00% (0.00-60.24%). n=4<br>3-7: 25.00% (3.19-65.09%). n=8<br>8-13: 86.36% (65.09-97.09%). n=22<br>≥14:100.00% (95.89-100.00%). n=88 | *Days after first PCR-positivity*<br>0-6: 61.05% (50.50-70.89%). n=95<br>7-13: 97.50% (92.87-99.48%). n=120<br>≥14:100.00% (92.45-100.00%). n=47 | *Days after first PCR-positivity*<br>0-6: 53.49% (42.41-64.23%). n=86<br>7-13: 93.44% (84.05-98.18%). n=61<br>≥14:100.00% (91.59-100.00%). n=42 |
| **Accuracy** | Within-day: 1.3-2.8%<br>Between-day: 2.2-7.2% | Within-day: 1.1-5.9%<br>Between-day: 1.2-5.9% | Within-day: 2.6-10.8%<br>Between-day: 5.2-12.4% | Within-day: 2.0-4.6%<br>Between-day: 2.8-6.1% |

In brackets 95% confidence intervals are shown. Abbreviations: CE-IVD, European Conformity - In Vitro Diagnostic Medical Device; CLIA, chemiluminescence immunoassay; CMIA, chemiluminescent microparticle immunoassay; ECLIA, electrochemiluminiscence immunoassay; ELISA, enzyme-linked immunosorbent assay; EUA: Emergency Use Authorization; K, potassium; Li, Lithium; n: number of samples tested; RBD: receptor-binding domain; S/Co, signal-to-cutoff ratio. [a] indicates the platform used in our study.

#### **Supplemental Table S2.** Diagnostic specificity of the four assays.

| Assay | Total | Negative | Positive | Specificity |
|---|---|---|---|---|
| Roche Elecsys Anti-SARS-CoV-2 | 341 | 340 | 1 | 99.71% (98.38-99.99%) |
| Abbott SARS-CoV-2 IgG | 298 | 296 | 2 | 99.33% (97.60-99.92%) |
| Siemens SARS-CoV-2 total (COV2T) | 288 | 287 | 1 | 99.65% (98.08-99.99%) |
| Siemens SARS-CoV-2 IgG (COV2G) | 191 | 191 | 0 | 100.00% (98.09-100.00%) |

Specificity of the assays was determined on 350 pre-pandemic samples; 95% confidence intervals are given in brackets.

**Supplemental Table S3.** False positive results in pre-pandemic samples.

| Sample number | Roche Elecsys Anti-SARS-CoV-2 | | Abbott SARS-CoV-2 IgG | | Siemens SARS-CoV-2 total (COV2T) | | Siemens SARS-CoV-2 IgG (COV2G) | |
|---|---|---|---|---|---|---|---|---|
| 1 | n | 0.08 | p | 1.46 | n | 0.13 | | |
| 2 | p | 1.18 | n | 0.04 | | | | |
| 3 | n | 0.09 | p | 2.42 | n | 0.21 | n | 0.17 |
| 4 | n | 0.09 | | | p | 2.91 | n | 0.04 |

Individual results (assay index) of all samples tested positive in at least one assay of 350 pre-pandemic samples are shown. Positive results (p) are shown in red, negative results (n) are shown in green.

**Supplemental Table S4.** Measurements of the immunoglobulin formulation Privigen®, undiluted and 1:50 diluted.

| Privigen | Roche Elecsys Anti-SARS-CoV-2 | Abbott SARS-CoV-2 IgG | Siemens SARS-CoV-2 total (COV2T) | Siemens SARS-CoV-2 IgG (COV2G) |
|---|---|---|---|---|
| Undiluted | 0.14 | 0.65 | <0.05 | 1.09 |
| 1:50, negative serum | 0.08 | 0.01 | <0.05 | 0.01 |
| 1:50, sodium chloride | 0.15 | 0.05 | 0.26 | 0.23 |

Data indicate the respective results of the raw data indexes of the tests. Privigen® was diluted with negative pre-pandemic serum or sodium chloride, respectively.

**Supplemental Table S5.** Individual results of all 230 sensitivity samples.

| # | # | Disease course | Days | Roche Elecsys Anti-SARS-CoV-2 | | Abbott SARS-CoV-2 IgG | | Siemens SARS-CoV-2 total (COV2T) | | Siemens SARS-CoV-2 IgG (COV2G) | |
|---|---|---|---|---|---|---|---|---|---|---|---|
| 1 | 1 | outpatient | 16 | p | 4.42 | p | 4.81 | | | | |
| 2 | 2 | outpatient | 20 | p | 4.46 | p | 4.24 | | | | |
| 3 | 3 | outpatient | 21 | p | 19.93 | p | 6.92 | p | 6.50 | p | 3.45 |
| 4 | 4 | outpatient | 24 | p | 24.56 | p | 6.72 | p | >10.00 | p | 3.20 |
| 5 | 5 | outpatient | 25 | p | 17.66 | p | 1.49 | | | | |
| 6 | 6 | outpatient | 25 | p | 5.32 | p | 3.85 | | | | |
| 7 | 7 | outpatient | 27 | p | 26.00 | p | 4.80 | p | >10.00 | p | 1.96 |
| 8 | 8 | outpatient | 30 | p | 3.82 | p | 3.05 | p | 1.58 | n | 0.32 |
| 9 | 9 | outpatient | 30 | p | 46.16 | p | 9.15 | | | | |
| 10 | 10 | outpatient | 32 | p | 2.86 | n | 0.71 | p | 4.07 | p | 1.28 |
| 11 | 11 | outpatient | 32 | p | 3.00 | p | 1.78 | p | 6.14 | p | 2.59 |
| 12 | 12 | outpatient | 33 | p | 30.01 | p | 4.68 | p | 3.63 | n | 0.36 |
| 13 | 13 | outpatient | 34 | n | 0.24 | n | 0.03 | n | 0.29 | n | 0.00 |
| 14 | 14 | outpatient | 36 | p | 28.47 | p | 4.96 | p | 4.84 | p | 2.03 |
| 15 | 15 | outpatient | 38 | p | 8.58 | p | 3.10 | p | 2.55 | p | 1.14 |
| 16 | 16 | outpatient | 38 | n | 0.11 | n | 0.01 | | | n | 0.06 |
| 17 | 17 | outpatient | 39 | p | 38.64 | p | 7.68 | p | >10.00 | p | 11.35 |
| 18 | 18 | outpatient | 39 | p | 43.54 | p | 5.66 | p | 3.86 | p | 1.97 |
| 19 | 19 | outpatient | 40 | p | 13.36 | p | 2.45 | p | >10.00 | p | 14.28 |
| 20 | 20 | outpatient | 40 | p | 84.46 | p | 7.99 | p | >10.00 | p | 12.05 |
| 21 | 21 | outpatient | 42 | p | 4.58 | p | 2.52 | | | | |

| | | | | | | | | | | | |
|---|---|---|---|---|---|---|---|---|---|---|---|
| 22 | 22 | outpatient | 42 | p | 32.47 | p | 4.04 | | | | |
| 23 | 23 | outpatient | 43 | p | 88.73 | p | 8.67 | p | >10.00 | p | 6.75 |
| 24 | 24 | outpatient | 43 | n | 0.11 | n | 0.18 | n | <0.05 | n | 0.04 |
| 25 | 25 | outpatient | 45 | p | 92.52 | p | 6.79 | p | 1.98 | p | 1.86 |
| 26 | 26 | outpatient | 45 | p | 39.48 | p | 9.90 | p | 7.36 | p | 6.76 |
| 27 | 27 | outpatient | 46 | p | 108.40 | p | 7.56 | p | 9.68 | p | 1.11 |
| 28 | 28 | outpatient | 47 | p | 14.35 | p | 2.29 | p | 2.48 | n | 0.15 |
| 29 | 29 | outpatient | 47 | p | 144.40 | p | 3.94 | p | 9.11 | p | 1.80 |
| 30 | 30 | outpatient | 48 | p | 58.86 | p | 6.18 | p | >10.00 | p | 9.86 |
| 31 | 31 | outpatient | 48 | p | 9.34 | p | 2.64 | p | 3.12 | n | 0.39 |
| 32 | 32 | outpatient | 48 | p | 31.36 | p | 5.13 | p | 3.38 | n | 0.50 |
| 33 | 33 | outpatient | 48 | p | 68.75 | p | 6.04 | p | 1.35 | p | 1.08 |
| 34 | 34 | outpatient | 49 | p | 29.76 | p | 2.31 | p | >10.00 | p | 1.86 |
| 35 | 35 | outpatient | 49 | p | 33.96 | p | 4.70 | n | 0.87 | n | 0.45 |
| 36 | 36 | outpatient | 49 | n | 0.11 | n | 0.30 | n | 0.28 | n | 0.10 |
| 37 | 37 | outpatient | 50 | p | 47.36 | p | 6.32 | p | 7.65 | p | 1.86 |
| 38 | 38 | outpatient | 50 | p | 27.04 | p | 3.93 | p | 7.76 | n | 0.62 |
| 39 | 39 | outpatient | 51 | p | 67.02 | p | 6.94 | p | >10.00 | p | 5.97 |
| 40 | 40 | outpatient | 51 | p | 69.06 | p | 9.11 | p | 9.79 | p | 2.53 |
| 41 | 41 | outpatient | 52 | p | 37.19 | p | 6.63 | p | 5.94 | n | 0.88 |
| 42 | 42 | outpatient | 52 | p | 12.66 | p | 3.18 | p | 2.33 | n | 0.55 |
| 43 | 43 | outpatient | 53 | p | 45.49 | p | 4.17 | p | 3.50 | p | 1.45 |
| 44 | 44 | outpatient | 55 | n | 0.09 | n | 0.07 | n | 0.40 | n | 0.12 |
| 45 | 45 | outpatient | 55 | p | 198.00 | p | 5.73 | p | 8.16 | p | 2.84 |
| 46 | 46 | outpatient | 56 | p | 20.74 | n | 1.23 | | | n | 0.37 |
| 47 | 47 | outpatient | 57 | p | 85.05 | p | 9.21 | p | >10.00 | p | 16.47 |
| 48 | 48 | outpatient | 58 | p | 69.65 | p | 5.16 | p | 1.08 | n | 0.22 |
| 49 | 49 | outpatient | 58 | p | 88.64 | p | 9.55 | p | >10.00 | p | 1.37 |
| 50 | 50 | outpatient | 58 | p | 49.01 | p | 5.58 | p | 3.62 | n | 0.64 |
| 51 | 51 | outpatient | 59 | p | 50.49 | p | 4.64 | p | >10.00 | p | 1.34 |
| 52 | 52 | outpatient | 59 | p | 37.06 | p | 4.50 | p | 4.45 | p | 1.67 |
| 53 | 53 | outpatient | 59 | p | 68.29 | p | 5.98 | p | >10.00 | p | 3.51 |
| 54 | 54 | outpatient | 60 | p | 7.79 | p | 2.66 | p | 2.86 | n | 0.53 |
| 55 | 55 | outpatient | 61 | p | 13.53 | p | 1.53 | p | 2.59 | p | 1.34 |
| 56 | 56 | outpatient | 61 | p | 45.69 | p | 4.23 | n | 0.27 | n | 0.28 |
| 57 | 57 | outpatient | 61 | p | 77.00 | p | 8.41 | p | >10.00 | p | >20.00 |
| 58 | 58 | outpatient | 62 | p | 229.00 | p | 9.10 | p | >10.00 | p | 5.83 |
| 59 | 59 | outpatient | 64 | p | 23.78 | p | 7.16 | p | 1.95 | p | 3.73 |
| 60 | 60 | outpatient | 64 | p | 24.67 | p | 4.25 | p | 5.82 | n | 0.44 |
| 61 | 61 | outpatient | 65 | p | 122.10 | p | 6.40 | p | 9.17 | p | 1.03 |
| 62 | 62 | outpatient | 65 | p | 51.98 | p | 5.67 | p | 5.97 | n | 0.57 |
| 63 | 63 | outpatient | 65 | p | 3.15 | n | 0.70 | p | 1.54 | n | 0.21 |
| 64 | 64 | outpatient | 66 | p | 15.00 | p | 2.79 | p | >10.00 | p | 1.12 |
| 65 | 65 | outpatient | 68 | n | 0.09 | n | 0.02 | n | 0.13 | n | 0.07 |
| 66 | 66 | outpatient | 69 | p | 28.61 | p | 2.16 | p | 7.83 | p | 2.17 |
| 67 | 67 | outpatient | 70 | p | 1.52 | n | 0.84 | p | 1.27 | n | 0.22 |
| 68 | 68 | outpatient | 71 | p | 209.50 | p | 6.20 | p | 9.04 | p | 2.00 |

| | | | | | | | | | | | |
|---|---|---|---|---|---|---|---|---|---|---|---|
| 69 | 69 | outpatient | 71 | p | 97.84 | p | 3.59 | p | 9.45 | p | 2.53 |
| 70 | 70 | outpatient | 72 | p | 53.74 | p | 3.54 | p | >10.00 | p | 2.09 |
| 71 | 71 | outpatient | 72 | p | 13.01 | n | 0.87 | p | 5.62 | p | 1.15 |
| 72 | 72 | outpatient | 75 | n | 0.11 | n | 0.02 | n | 0.11 | n | 0.03 |
| 73 | 73 | outpatient | 77 | p | 18.45 | p | 3.46 | p | 6.68 | p | 1.14 |
| 74 | 74 | outpatient | 77 | p | 149.40 | p | 9.26 | p | >10.00 | p | 12.18 |
| 75 | 75 | outpatient | 82 | p | 215.30 | p | 3.58 | p | >10.00 | p | 1.12 |
| 76 | 76 | outpatient | 82 | p | 163.90 | p | 7.97 | | | | |
| 77 | 77 | outpatient | 85 | p | 159.60 | p | 6.16 | p | >10.00 | p | 2.38 |
| 78 | 78 | outpatient | 87 | p | 126.50 | p | 3.04 | p | 5.54 | n | 0.89 |
| 79 | 79 | outpatient | 89 | p | 159.00 | p | 2.73 | p | >10.00 | p | 1.12 |
| 80 | 80 | outpatient | 90 | p | 153.00 | p | 6.37 | p | 6.93 | p | 3.95 |
| 81 | 81 | outpatient | 90 | p | 212.20 | p | 7.67 | p | 4.28 | p | 1.31 |
| 82 | 82 | outpatient | 97 | p | 194.20 | p | 4.53 | p | >10.00 | p | 6.43 |
| 83 | 83 | outpatient | 104 | p | 51.14 | p | 2.06 | | | | |
| 84 | 84 | outpatient | 111 | p | 72.73 | p | 3.70 | | | | |
| 85 | 85 | outpatient | 120 | p | 93.09 | p | 3.61 | | | | |
| 86 | 1 | general ward | 14 | p | 15.79 | p | 4.55 | p | >10.00 | p | 7.58 |
| 87 | 2 | general ward | 14 | p | 8.29 | p | 5.80 | p | 8.52 | p | 1.74 |
| 88 | 3 | general ward | 14 | p | 8.53 | p | 6.61 | p | 4.99 | p | 17.86 |
| 89 | 4 | general ward | 14 | n | 0.47 | n | 0.94 | n | 0.34 | n | 0.12 |
| 90 | 5 | general ward | 14 | p | 3.56 | p | 3.14 | p | >10.00 | n | 0.45 |
| 91 | 6 | general ward | 14 | p | 3.25 | p | 6.66 | | | | |
| 92 | 7 | general ward | 14 | p | 5.27 | p | 4.02 | | | | |
| 93 | 8 | general ward | 15 | p | 10.91 | p | 3.63 | p | >10.00 | p | 5.67 |
| 94 | 9 | general ward | 15 | p | 27.00 | p | 4.74 | p | 9.13 | p | 2.47 |
| 95 | 10 | general ward | 15 | p | 15.63 | p | 4.70 | p | 3.42 | n | 0.24 |
| 96 | 11 | general ward | 15 | p | 24.61 | p | 8.39 | p | >10.00 | p | 10.50 |
| 97 | 12 | general ward | 15 | p | 45.55 | p | 8.27 | p | >10.00 | p | >20.00 |
| 98 | 13 | general ward | 15 | p | 1.65 | p | 3.93 | | | | |
| 99 | 14 | general ward | 15 | p | 13.66 | p | 1.87 | | | | |
| 100 | 15 | general ward | 16 | p | 14.70 | p | 8.47 | p | >10.00 | p | 14.47 |
| 101 | 16 | general ward | 16 | p | 11.99 | p | 6.45 | p | >10.00 | p | 9.65 |
| 102 | 17 | general ward | 16 | p | 18.97 | n | 0.78 | p | >10.00 | p | >20.00 |
| 103 | 18 | general ward | 17 | p | 3.50 | p | 5.96 | p | >10.00 | p | 1.85 |
| 104 | 19 | general ward | 17 | p | 3.71 | p | 5.69 | p | >10.00 | p | 7.32 |
| 105 | 20 | general ward | 17 | p | 14.37 | p | 8.71 | p | 2.55 | p | 3.42 |
| 106 | 21 | general ward | 17 | p | 52.97 | p | 8.96 | p | >10.00 | p | >20.00 |
| 107 | 22 | general ward | 17 | p | 25.00 | p | 7.05 | p | 7.06 | p | 11.61 |
| 108 | 23 | general ward | 17 | p | 15.84 | p | 4.97 | | | | |
| 109 | 24 | general ward | 17 | p | 1.43 | | | | | | |
| 110 | 25 | general ward | 18 | p | 19.79 | p | 2.08 | p | >10.00 | p | 2.66 |
| 111 | 26 | general ward | 18 | p | 11.51 | p | 7.59 | p | >10.00 | p | 16.18 |
| 112 | 27 | general ward | 18 | p | 21.65 | p | 8.25 | p | >10.00 | p | 14.52 |
| 113 | 28 | general ward | 19 | p | 19.30 | p | 8.08 | p | >10.00 | p | 15.71 |
| 114 | 29 | general ward | 19 | n | 0.25 | p | 8.30 | n | 0.67 | n | 0.03 |
| 115 | 30 | general ward | 19 | n | 0.18 | n | 0.08 | p | 1.75 | n | 0.53 |

| | | | | | | | | | | | |
|---|---|---|---|---|---|---|---|---|---|---|---|
| 116 | 31 | general ward | 19 | p | 9.95 | p | 5.98 | p | >10.00 | p | 6.80 |
| 117 | 32 | general ward | 19 | p | 37.50 | p | 6.12 | p | >10.00 | p | >20.00 |
| 118 | 33 | general ward | 20 | p | 4.02 | p | 3.38 | p | >10.00 | p | 8.24 |
| 119 | 34 | general ward | 20 | p | 22.21 | p | 7.05 | p | >10.00 | p | >20.00 |
| 120 | 35 | general ward | 20 | p | 5.84 | p | 1.84 | n | 0.68 | n | 0.29 |
| 121 | 36 | general ward | 20 | p | 13.03 | p | 6.69 | | | p | 3.80 |
| 122 | 37 | general ward | 20 | p | 9.58 | p | 4.86 | | | | |
| 123 | 38 | general ward | 21 | p | 1.15 | p | 5.28 | n | 0.29 | n | 0.50 |
| 124 | 39 | general ward | 21 | p | 36.54 | p | 7.18 | p | >10.00 | p | 16.86 |
| 125 | 40 | general ward | 21 | p | 21.26 | p | 6.13 | p | >10.00 | p | >20.00 |
| 126 | 41 | general ward | 21 | p | 13.90 | p | 5.50 | p | >10.00 | p | 11.43 |
| 127 | 42 | general ward | 22 | p | 39.78 | p | 8.90 | p | >10.00 | p | 5.74 |
| 128 | 43 | general ward | 23 | p | 33.15 | p | 8.25 | p | >10.00 | p | >20.00 |
| 129 | 44 | general ward | 23 | p | 21.92 | p | 8.16 | p | >10.00 | p | >20.00 |
| 130 | 45 | general ward | 24 | p | 44.42 | p | 2.25 | p | 5.83 | p | 1.68 |
| 131 | 46 | general ward | 24 | n | 0.80 | p | 6.47 | p | >10.00 | p | 10.70 |
| 132 | 47 | general ward | 24 | p | 14.02 | p | 7.35 | p | >10.00 | p | 14.59 |
| 133 | 48 | general ward | 24 | p | 39.26 | p | 9.46 | p | >10.00 | p | 6.44 |
| 134 | 49 | general ward | 24 | p | 41.21 | p | 8.50 | p | >10.00 | p | >20.00 |
| 135 | 50 | general ward | 25 | p | 35.27 | p | 8.70 | p | >10.00 | p | >20.00 |
| 136 | 51 | general ward | 26 | p | 9.41 | p | 6.88 | p | >10.00 | p | >20.00 |
| 137 | 52 | general ward | 27 | p | 12.94 | p | 8.00 | p | >10.00 | p | 9.12 |
| 138 | 53 | general ward | 28 | p | 15.53 | p | 8.83 | p | >10.00 | p | 6.44 |
| 139 | 54 | general ward | 28 | p | 3.23 | p | 5.91 | p | >10.00 | p | >20.00 |
| 140 | 55 | general ward | 28 | p | 35.48 | p | 8.77 | p | >10.00 | p | 8.24 |
| 141 | 56 | general ward | 33 | p | 60.67 | p | 6.60 | | | p | 14.07 |
| 142 | 57 | general ward | 34 | p | 84.65 | p | 8.00 | p | >10.00 | | |
| 143 | 58 | general ward | 39 | p | 70.82 | p | 9.50 | | | | |
| 144 | 59 | general ward | 44 | p | 86.18 | p | 8.53 | p | >10.00 | p | 4.46 |
| 145 | 60 | general ward | 46 | p | 87.75 | p | 6.27 | p | >10.00 | p | 5.15 |
| 146 | 61 | general ward | 46 | p | 53.67 | p | 7.24 | p | >10.00 | p | 4.82 |
| 147 | 62 | general ward | 46 | p | 95.25 | p | 9.12 | | | | |
| 148 | 63 | general ward | 47 | p | 75.33 | p | 6.05 | p | >10.00 | p | 5.16 |
| 149 | 64 | general ward | 48 | p | 64.57 | p | 7.68 | p | >10.00 | p | 16.59 |
| 150 | 65 | general ward | 49 | p | 14.45 | p | 4.60 | p | >10.00 | p | 2.60 |
| 151 | 66 | general ward | 50 | n | 0.09 | n | 0.01 | n | 0.19 | n | 0.03 |
| 152 | 67 | general ward | 50 | p | 63.92 | p | 6.15 | p | >10.00 | p | >20.00 |
| 153 | 68 | general ward | 50 | n | 0.32 | n | 0.69 | n | 0.10 | n | 0.93 |
| 154 | 69 | general ward | 51 | p | 51.54 | p | 8.63 | p | >10.00 | p | 6.90 |
| 155 | 70 | general ward | 51 | p | 47.70 | p | 6.30 | p | 2.73 | p | 1.01 |
| 156 | 71 | general ward | 53 | p | 15.11 | p | 7.84 | n | 0.75 | p | 1.43 |
| 157 | 72 | general ward | 54 | p | 19.18 | p | 8.44 | p | >10.00 | p | 7.18 |
| 158 | 73 | general ward | 54 | p | 66.68 | p | 8.18 | p | >10.00 | p | 4.95 |
| 159 | 74 | general ward | 57 | p | 76.02 | p | 7.48 | p | >10.00 | p | 4.90 |
| 160 | 75 | general ward | 58 | p | 70.55 | p | 7.95 | p | >10.00 | p | >20.00 |
| 161 | 76 | general ward | 58 | p | 3.94 | n | 1.35 | p | 2.78 | n | 0.54 |
| 162 | 77 | general ward | 58 | p | 36.85 | p | 3.64 | | | | |

| | | | | | | | | | | | |
|---|---|---|---|---|---|---|---|---|---|---|---|
| 163 | 78 | general ward | 59 | p | 50.29 | p | 2.53 | n | 0.36 | n | 0.55 |
| 164 | 79 | general ward | 59 | p | 98.70 | p | 9.02 | p | >10.00 | p | 3.13 |
| 165 | 80 | general ward | 59 | p | 42.36 | p | 7.14 | p | >10.00 | p | >20.00 |
| 166 | 81 | general ward | 60 | p | 53.45 | p | 7.65 | | | | |
| 167 | 82 | general ward | 61 | p | 49.65 | p | 6.58 | p | >10.00 | p | 7.04 |
| 168 | 83 | general ward | 61 | p | 210.10 | p | 7.34 | p | 9.51 | p | 1.97 |
| 169 | 84 | general ward | 61 | p | 64.63 | p | 3.42 | p | >10.00 | | |
| 170 | 85 | general ward | 62 | p | 92.64 | p | 7.43 | p | >10.00 | p | 8.49 |
| 171 | 86 | general ward | 62 | p | 16.23 | p | 2.50 | p | 7.66 | p | 2.05 |
| 172 | 87 | general ward | 63 | n | 0.10 | n | 0.04 | n | <0.05 | n | 0.07 |
| 173 | 88 | general ward | 63 | p | 84.20 | p | 9.44 | p | >10.00 | p | 7.20 |
| 174 | 89 | general ward | 63 | p | 60.58 | p | 9.03 | p | >10.00 | p | 6.47 |
| 175 | 90 | general ward | 63 | p | 94.06 | p | 8.45 | p | >10.00 | p | >20.00 |
| 176 | 91 | general ward | 65 | p | 161.80 | p | 8.39 | p | 5.91 | p | 2.84 |
| 177 | 92 | general ward | 65 | p | 139.40 | p | 7.02 | p | >10.00 | p | 4.39 |
| 178 | 93 | general ward | 67 | p | 70.86 | p | 5.13 | p | >10.00 | p | 4.91 |
| 179 | 94 | general ward | 68 | p | 36.67 | p | 3.42 | p | 2.87 | n | 0.87 |
| 180 | 95 | general ward | 69 | p | 71.77 | p | 9.45 | p | >10.00 | p | 4.72 |
| 181 | 96 | general ward | 70 | p | 80.46 | p | 5.26 | p | 1.87 | n | 0.33 |
| 182 | 97 | general ward | 71 | p | 81.51 | p | 4.53 | p | >10.00 | p | >20.00 |
| 183 | 98 | general ward | 71 | p | 92.71 | p | 9.59 | p | 5.57 | p | 2.79 |
| 184 | 99 | general ward | 71 | p | 131.20 | p | 7.04 | p | >10.00 | p | 10.69 |
| 185 | 100 | general ward | 71 | p | 141.40 | p | 8.91 | p | >10.00 | p | 11.69 |
| 186 | 101 | general ward | 72 | p | 178.50 | p | 7.33 | p | >10.00 | p | 19.26 |
| 187 | 102 | general ward | 73 | p | 85.78 | p | 9.28 | p | >10.00 | | |
| 188 | 103 | general ward | 74 | p | 71.67 | p | 8.98 | p | >10.00 | p | 18.08 |
| 189 | 104 | general ward | 96 | p | 104.10 | p | 7.02 | | | | |
| 190 | 105 | general ward | 107 | p | 106.40 | p | 7.50 | | | | |
| 191 | 106 | general ward | 108 | p | 17.44 | p | 1.56 | | | | |
| 192 | 1 | intensive care | 18 | p | 17.25 | p | 7.01 | p | >10.00 | p | 16.86 |
| 193 | 2 | intensive care | 19 | p | 68.42 | p | 8.86 | p | >10.00 | p | 7.03 |
| 194 | 3 | intensive care | 21 | p | 26.31 | p | 8.42 | p | >10.00 | p | >20.00 |
| 195 | 4 | intensive care | 21 | p | 9.31 | p | 6.68 | p | >10.00 | p | 10.95 |
| 196 | 5 | intensive care | 21 | p | 43.80 | p | 8.02 | p | >10.00 | p | 3.41 |
| 197 | 6 | intensive care | 21 | p | 47.57 | p | 4.40 | | | | |
| 198 | 7 | intensive care | 23 | p | 1.35 | p | 7.65 | p | >10.00 | p | 1.86 |
| 199 | 8 | intensive care | 24 | p | 52.95 | p | 9.54 | p | >10.00 | p | >20.00 |
| 200 | 9 | intensive care | 24 | p | 47.73 | p | 8.03 | p | >10.00 | p | >20.00 |
| 201 | 10 | intensive care | 25 | p | 75.09 | p | 8.76 | p | >10.00 | p | 7.59 |
| 202 | 11 | intensive care | 26 | p | 28.22 | p | 8.70 | p | >10.00 | p | >20.00 |
| 203 | 12 | intensive care | 26 | p | 77.10 | p | 8.63 | p | >10.00 | p | >20.00 |
| 204 | 13 | intensive care | 27 | p | 11.43 | p | 7.59 | p | >10.00 | p | 8.59 |
| 205 | 14 | intensive care | 27 | p | 38.86 | p | 8.60 | p | >10.00 | p | 14.35 |
| 206 | 15 | intensive care | 28 | p | 5.54 | p | 7.70 | p | >10.00 | p | 5.21 |
| 207 | 16 | intensive care | 28 | p | 10.13 | p | 6.24 | p | >10.00 | p | 7.25 |
| 208 | 17 | intensive care | 28 | p | 39.27 | p | 7.61 | p | >10.00 | p | >20.00 |
| 209 | 18 | intensive care | 28 | p | 7.43 | n | 1.36 | p | 2.74 | | |

| | | | | | | | | | | | |
|---|---|---|---|---|---|---|---|---|---|---|---|
| 210 | 19 | intensive care | 28 | p | 6.42 | p | 4.99 | p | >10.00 | p | >20.00 |
| 211 | 20 | intensive care | 28 | p | 52.77 | p | 8.17 | p | >10.00 | p | 13.39 |
| 212 | 21 | intensive care | 28 | p | 23.44 | p | 8.40 | p | >10.00 | p | 19.09 |
| 213 | 22 | intensive care | 29 | p | 75.52 | p | 9.26 | p | >10.00 | p | >20.00 |
| 214 | 23 | intensive care | 31 | p | 16.55 | p | 8.30 | p | >10.00 | p | 6.73 |
| 215 | 24 | intensive care | 31 | p | 70.01 | p | 8.76 | p | >10.00 | p | >20.00 |
| 216 | 25 | intensive care | 31 | p | 72.69 | p | 8.99 | p | >10.00 | p | >20.00 |
| 217 | 26 | intensive care | 31 | p | 77.61 | p | 8.34 | p | >10.00 | p | >20.00 |
| 218 | 27 | intensive care | 34 | n | 0.08 | n | 0.01 | n | 0.20 | n | 0.13 |
| 219 | 28 | intensive care | 35 | n | 0.11 | | | n | <0.05 | | |
| 220 | 29 | intensive care | 40 | p | 77.42 | p | 9.29 | | | | |
| 221 | 30 | intensive care | 41 | p | 58.18 | p | 7.68 | p | >10.00 | p | 7.46 |
| 222 | 31 | intensive care | 65 | p | 44.20 | p | 4.77 | | | | |
| 223 | 32 | intensive care | 66 | p | 117.50 | p | 8.01 | p | >10.00 | p | 19.63 |
| 224 | 33 | intensive care | 67 | p | 191.60 | p | 8.63 | p | >10.00 | p | 9.48 |
| 225 | 34 | intensive care | 78 | p | 94.79 | p | 4.24 | p | >10.00 | p | >20.00 |
| 226 | 35 | intensive care | 80 | p | 84.76 | p | 6.06 | p | >10.00 | | |
| 227 | 36 | intensive care | 84 | p | 88.00 | p | 8.24 | p | >10.00 | p | >20.00 |
| 228 | 37 | intensive care | 87 | p | | p | 8.76 | | | | |
| 229 | 38 | intensive care | 98 | p | 105.70 | p | 8.90 | | | | |
| 230 | 39 | intensive care | 99 | p | 73.59 | p | 8.74 | | | | |

Individual results (raw data assay index) of the samples of SARS-CoV-2 PCR-confirmed patients are shown (one sample per patient; the sample ≥14 days and closest to day 28 after symptom onset was used). Positive results (p) are shown in red, negative results (n) are shown in green. Days indicate the time between disease onset and blood sampling. # indicates the patient number.

**Supplemental Table S6.** Diagnostic sensitivity of the four assays stratified for disease course, age and gender.

| | | Roche Elecsys Anti-SARS-CoV-2 | Abbott SARS-CoV-2 IgG | Siemens SARS-CoV-2 total (COV2T) | Siemens SARS-CoV-2 IgG (COV2G) |
|---|---|---|---|---|---|
| **Sensitivity according to PI** | | **99.50% (97.00-100.00%)** | **100.00% (95.89-100.00%)** | **100.00% (92.45-100.00%)** | **100.00% (91.59-100.00%)** |
| **All patients** | Total | 230 | 229 | 195 | 193 |
| | Positive | 214 | 208 | 176 | 152 |
| | Negative | 16 | 21 | 19 | 41 |
| | Sensitivity | 93.04% (88.95-95.97%) | 90.83% (86.32-94.23%) | 90.26% (85.20-94.03%) | 78.76% (72.30-84.30%) |
| **Outpatient** | Total | 85 | 85 | 72 | 74 |
| | Positive | 78 | 73 | 64 | 48 |
| | Negative | 7 | 12 | 8 | 26 |
| | Sensitivity | 91.76% (83.77-96.62%) | 85.88% (76.64-92.49%) | 88.89% (79.28-95.08%) | 64.86% (52.89-75.61%) |
| **Gen. ward** | Total | 106 | 105 | 90 | 89 |
| | Positive | 99 | 98 | 81 | 75 |
| | Negative | 7 | 7 | 9 | 14 |
| | Sensitivity | 93.40% (86.87-97.30%) | 93.33% (86.75-97.28%) | 90.00% (81.86-95.32%) | 84.27% (75.02-91.12%) |
| **ICU** | Total | 39 | 38 | 33 | 30 |
| | Positive | 37 | 36 | 31 | 29 |
| | Negative | 2 | 2 | 2 | 1 |
| | Sensitivity | 94.87% (82.68-99.37%) | 94.74% (82.25-99.36%) | 93.94% (79.77-99.26%) | 96.67% (82.78-99.92%) |
| **18-49** | Total | 74 | 73 | 60 | 61 |
| | Positive | 70 | 63 | 53 | 39 |
| | Negative | 4 | 10 | 7 | 22 |
| | Sensitivity | 94.59% (86.73-98.51%) | 86.30% (76.25-93.23%) | 88.33% (77.43-95.18%) | 63.93% (50.63-75.84%) |
| **50-69** | Total | 109 | 109 | 97 | 95 |
| | Positive | 102 | 101 | 91 | 82 |
| | Negative | 7 | 8 | 6 | 13 |
| | Sensitivity | 93.58% (87.22-97.83%) | 92.66% (86.05-96.78%) | 93.81% (87.02-97.70%) | 86.32% (77.74-92.51%) |
| **70-100** | Total | 47 | 46 | 38 | 37 |
| | Positive | 42 | 43 | 32 | 31 |
| | Negative | 5 | 3 | 6 | 6 |
| | Sensitivity | 89.36% (76.90-96.45%) | 93.48% (82.10-98.63%) | 84.21% (68.75-93.98%) | 83.78% (67.99-93.81%) |
| Male | Total | 147 | 146 | 129 | 126 |
| | Positive | 140 | 138 | 120 | 110 |
| | Negative | 7 | 8 | 9 | 16 |
| | Sensitivity | 95.24% (90.43-98.06%) | 94.52% (89.49-97.60%) | 93.02% (87.17-96.76%) | 87.30% (80.20-92.56%) |
| Female | Total | 83 | 82 | 66 | 67 |
| | Positive | 74 | 69 | 56 | 42 |
| | Negative | 9 | 13 | 10 | 25 |
| | Sensitivity | 89.16% (80.41-94.92%) | 84.15% (74.42-91.28%) | 84.85% (73.90-92.49%) | 62.69% (50.01-74.20%) |

Sensitivity of the evaluated test-systems including all SARS-CoV-2 PCR-confirmed patients (one sample per patient; the sample ≥14 days and closest to day 28 after symptom onset was used) and stratified for severity of the disease (outpatient, hospitalised at the general ward, hospitalized at the intensive care unit) as well as age (18-49, 50-69, 70-100 years), respectively. 95% confidence intervals are given in brackets. Abbreviations: PI: package insert.

### **Supplemental Table S7.** Positivity rate across the time course of the disease.

| | Roche Elecsys Anti-SARS-CoV-2 | | Abbott SARS-CoV-2 IgG | | Siemens SARS-CoV-2 total (COV2T) | | Siemens SARS-CoV-2 IgG (COV2G) | |
|---|---|---|---|---|---|---|---|---|
| **days** | **n** | **Positivity rate** | **n** | **Positivity rate** | **n** | **Positivity rate** | **n** | **Positivity rate** |
| **0-6** | 43 | **25,58%** (13.25-41.17%) | 41 | **19,51%** (8.82-34.87%) | 28 | **32,14%** (15.88-52.35%) | 27 | **22,22%** (8.62-42.26%) |
| **7-13** | 114 | **52,63%** (43.06-62.06%) | 113 | **53,98%** (44.35-63.40%) | 97 | **54,64%** (44.21-64.78%) | 96 | **40,62%** (30.71-51.13%) |
| **14-20** | 107 | **91,59%** (84.63-96.08%) | 107 | **95,33%** (89.43-98.47%) | 95 | **88,42%** (80.23-94.08%) | 94 | **80,85%** (71.44-88.24%) |
| **21-40** | 122 | **90,98%** (84.44-95.41%) | 117 | **91,45%** (84.84-95.83%) | 109 | **90,83%** (83.77-95.51%) | 101 | **91,09%** (83.76-95.84%) |
| **>40** | 309 | **90,29%** (86.43-93.35%) | 306 | **89,87%** (85.93-93.01%) | 188 | **87,77%** (82.21-92.08%) | 180 | **77,78%** (70.99-83.62%) |

A total of 700 samples of 245 SARS-CoV-2 PCR-confirmed patients were measured. The table shows the positivity rate of the four investigated assays depending on the time course of the disease. Days indicate the time between disease onset and blood sampling. N indicates the total number of samples tested. 95% confidence intervals are given in brackets.

### **Supplemental Table S8.** ROC curve analysis of the four assays.

| **Assay** | **AUC** |
|---|---|
| Roche Elecsys Anti-SARS-CoV-2 | 0.984 (0.974-0.991) |
| Abbott SARS-CoV-2 IgG | 0.982 (0.970-0.990) |
| Siemens SARS-CoV-2 total (COV2T) | 0.975 (0.961-9.986) |
| Siemens SARS-CoV-2 IgG (COV2G) | 0.966 (0.948-0.980) |

ROC curve analysis was based on 350 pre-pandemic (=negative) samples (samples, Roche: 341, Abbott: 298, Siemens COV2T: 288, Siemens COV2G: 191) and the samples of SARS-CoV-2 PCR-confirmed (=positive) patients (samples, Roche: 537, Abbott: 530, Siemens COV2T: 392, Siemens COV2G: 375). Samples of SARS-CoV-2 positive patients were only considered if time-span between disease onset and blood draw was ≥14 days. 95% confidence intervals are given in brackets. Abbreviations: AUC, area under the curve; ROC, receiver operating characteristic.

**Supplemental Table S9.** Sensitivity and specificity of the assays using the modified cut-off index.

| | | Manufacturer's COI | Modified COI |
|---|---|---|---|
| | **COI** | **≥1.0: positive** | **>0.15: positive** |
| **Roche Elecsys Anti-SARS-CoV-2** | Sensitivity (n=230) | 93.04% (89.00-95.67%) | 95.65% (92.18-97.62%) |
| | Specificity (n=341) | 99.71% (98.36-99.95%) | 97.36% (95.06-98.61%) |
| | **COI** | **≥1.4: positive** | **>0.54: positive** |
| **Abbott SARS-CoV-2 IgG** | Sensitivity (n=228) | 90.79% (86.33-93.90%) | 95.18% (91.57-97.28%) |
| | Specificity (n=298) | 99.33% (97.59-99.82%) | 98.32% (96.13-99.28%) |
| | **COI** | **≥1.0: positive** | **>0.42: positive** |
| **Siemens SARS-CoV-2 total (COV2T)** | Sensitivity (n=195) | 90.26% (85.28-93.67%) | 92.31% (87.70-95.28%) |
| | Specificity (n=288) | 99.65% (98.06-99.94%) | 98.61% (96.48-99.46%) |
| | **COI** | **≥1.0: positive** | **>0.32: positive** |
| **Siemens SARS-CoV-2 IgG (COV2G)** | Sensitivity (n=193) | 78.76% (72.45-83.94%) | 89.64% (84.54-93.19%) |
| | Specificity (n=191) | 100.00% (98.03-100.00%) | 97.38% (94.02-98.88%) |

Sensitivity and specificity of the assays using different cut-off indexes (COI): the COI according to the manufacturer and a modified COI according to our ROC curve analysis. Sensitivity was evaluated on samples of SARS-CoV-2 PCR-confirmed patients (one sample per patient; the sample ≥14 days and closest to day 28 after symptom onset was used). Specificity was determined on 350 pre-pandemic samples. 95% confidence intervals are shown in brackets.

**Supplemental Table S10.** Concordance correlation coefficient between the four assays.

| Test | Roche | Abbott | Siemens COV2T | Siemens COV2G |
|---|---|---|---|---|
| **Roche** | x | 90.13 (n=972) | 92.49 (n=800) | 81.93 (n=689) |
| **Abbott** | | x | 85.47 (n=750) | 77.45 (n=645) |
| **Siemens COV2T** | | | x | 84.93 (n=680) |

Concordance correlation coefficient of the qualitative results of the four assays (350 pre-pandemic samples and all 700 samples of SARS-CoV-2 PCR-confirmed patients). Abbreviations: n, total number of samples measured with both assays. Abbott = Abbott SARS-CoV-2 IgG; Roche = Roche Elecsys Anti SARS-CoV-2; Siemens COV2G = Siemens SARS-CoV-2 IgG; Siemens COV2T = Siemens SARS-CoV-2 Total.

### Supplemental Figures

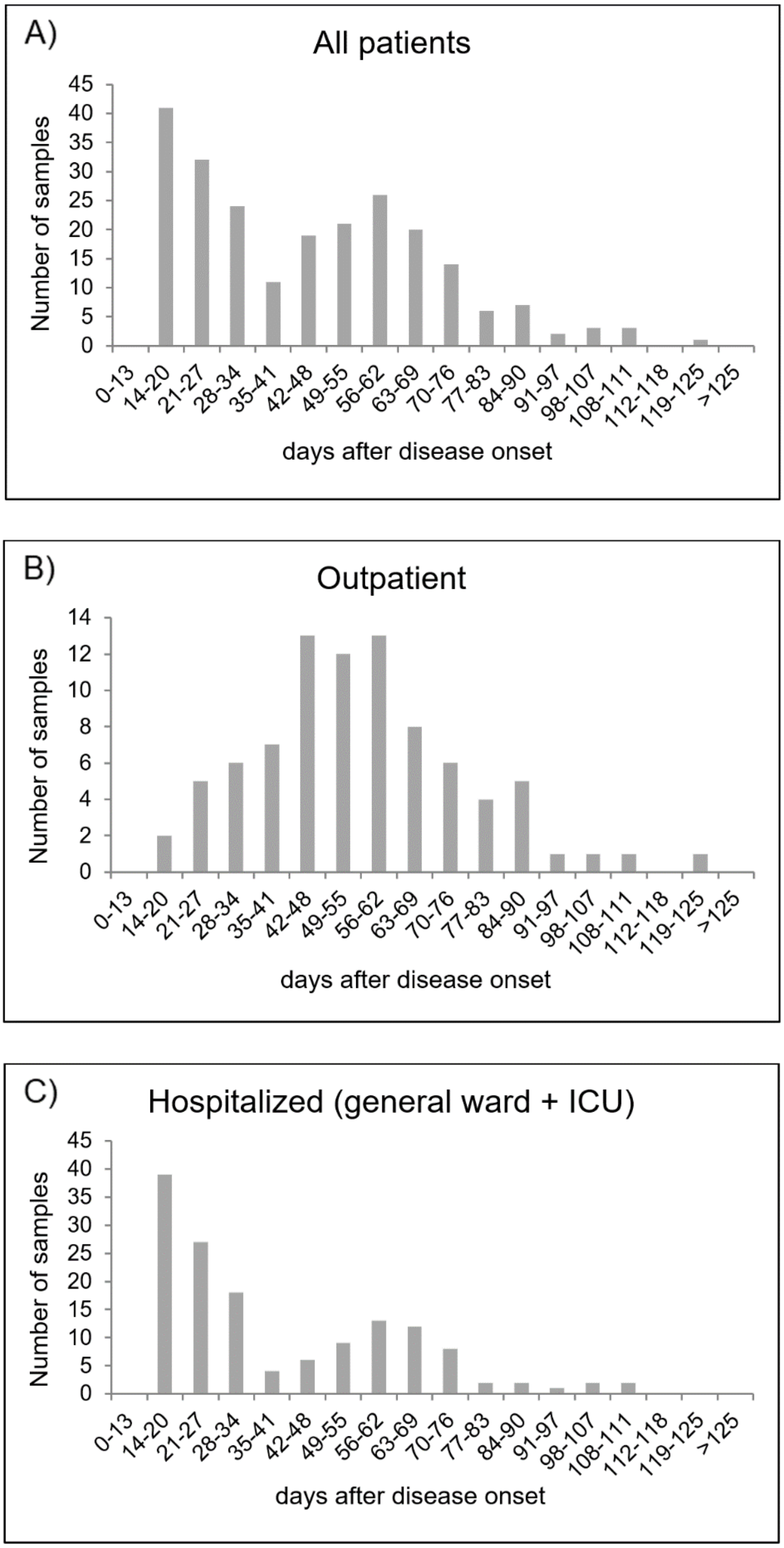


**Supplemental Figure S1. Distribution of sample timing.** Horizontal axis: time span between disease onset and timing of the blood draw of the sample used for the sensitivity analysis. Vertical axis: number of samples. A) all patients; B) outpatient; C) hospitalized patients (general ward and intensive care unit, ICU).

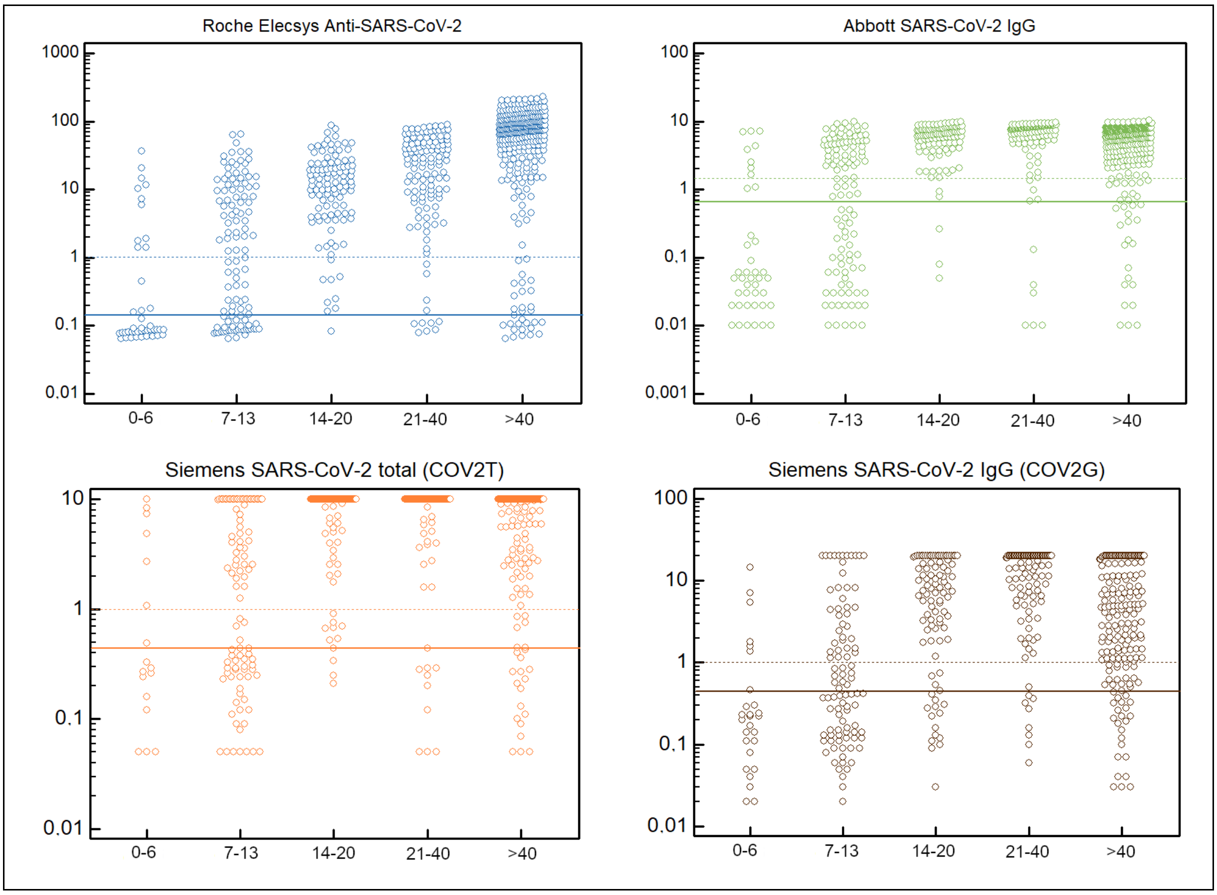


**Supplemental Figure S2. Raw data index results of all 700 samples of SARS-CoV-2 PCR-confirmed patients, stratified for the time course of the disease.** Horizontal axis: days after symptom onset. Vertical axis: raw data index of the respective test. Dashed horizontal line: manufacturer's cut-off index. Solid horizontal line: modified cut-off index.
